# An impactor-based aerosol platform for probing indoor, short-range transmission dynamics: a Phi6 bacteriophage study

**DOI:** 10.1101/2025.10.24.25338726

**Authors:** Mathura Thirugnanasampanthar, Mellissa Gomez, Ekaterina Kvitka, Kyle Jackson, Rod G Rhem, Myrna B Dolovich, Zeinab Hosseinidoust

## Abstract

1.

Short-range transmission is a driver of airborne disease spread. However, limited knowledge exists on the immediate impact of host, environmental, and seasonal factors on viable pathogen-laden droplets (VPLD) shortly after release. This work modelled the effects of respiratory droplet size, airway mucus composition, viral loads, and seasonal variations in indoor relative humidity (RH) on the number of VPLD collected shortly after equilibration. Clinically relevant concentrations of the SARS-CoV-2 surrogate bacteriophage, Phi6, were prepared in solutions reflecting the solute content of the airway mucus in healthy and disease states. Low solute levels and viral loads, characteristic of the presymptomatic phase, increased VPLD counts (initial diameter range, 5.59 to 20.28 µm) under low indoor RH in temperate winter. Elevated solute levels and decreased viral loads, characteristic of the late symptomatic phase, reduced VPLD counts (initial diameter range, 1.73 to 20.28 µm) under intermediate RH in temperate summers. High viral loads, characteristic of the early symptomatic phase, resulted in comparable VPLD counts (initial diameter range, 1.73 to 20.28 µm) across solute levels and RH, indicating a buffering effect of high viral loads against inactivation. These findings, integrated with complementary lines of evidence, were used to describe potential mechanisms of short-range transmission dynamics.

**SYNOPSIS:** This study investigated the effects of four factors including respiratory droplet size, mucus composition, viral load, and seasonal variations in indoor relative humidity, on the recovery of viable pathogen-laden droplets to understand short-range transmission dynamics.

## 4. INTRODUCTION

Nonpharmaceutical interventions (NPIs), including lockdowns, social distancing, travel bans, gathering restrictions, and the use of facemasks, were implemented during the early stages of the coronavirus disease 2019 (COVID-19) pandemic to mitigate the spread of severe acute respiratory syndrome coronavirus 2 (SARS-CoV-2).^1^ These measures were primarily based on the understanding that proximity to an infectious source heightens the risk of transmission.^2^ Moreover, evidence indicated that presymptomatic, asymptomatic, and mildly symptomatic carriers may be as infectious as highly symptomatic patients, making it challenging to implement selective quarantine measures.^3^ However, prolonged and indiscriminate enforcement of NPIs proved unsustainable, adversely impacting the psychological well-being of individuals and the socioeconomic stability of institutions worldwide.^4,5^ This highlighted an urgent need to unravel the transmission dynamics of this emerging respiratory pathogen.^6^

Airborne transmission has emerged as the primary driver of SARS-CoV-2 spread, mediated by infectious respiratory droplets generated during respiratory activities such as breathing, speaking, sneezing, coughing, laughing, and singing.^7,8^ This route is further classified into short-range or long-range transmission modes.^9^ Importantly, short-range transmission during face-to-face interactions is the most common mode of disease spread.^9,10,11,12^ Sustained interactions in enclosed spaces and contact with superspreaders significantly contributed to the rapid spread of SARS-CoV-2.^13^ Indoor environments with inadequate ventilation, stable relative humidity (RH) and temperature conditions, and limited ultraviolet radiation may slow the decay of pathogen viability, thereby increasing transmission risk.^14,15^ This is particularly concerning given that individuals in urban environments spend 90% of their time indoors.^16^

The composition of the airway mucus lining, shaped by genetic, age-related, and disease-specific factors, contributes to interindividual differences in respiratory tract physiology and may influence the transmissibility of SARS-CoV-2.^17,18,19,20,21^ Changes in airway mucus composition and viral load that occur before, during, and after symptom onset may provide insight into the transmission dynamics of individuals presenting with varying degrees of symptom severity.^10^ The impact of host, environmental, and seasonal factors on infectious respiratory droplets immediately after expulsion is needed to understand short-range transmission dynamics.

Experimental platforms are available for studying pathogen decay kinetics within different droplet size fractions over varying timescales.^22^ Goldberg rotating drums are widely used to study pathogen decay kinetics within polydisperse droplets over long timescales, ranging from hours to days.^22^ However, a limitation of Goldberg drum studies is their inability to perform endpoint droplet size measurements.^22^ As a result, these studies cannot pair pathogen viability changes to the physical transformation of the carrier droplet, which is critical for estimating airborne residence time and airway deposition efficiency.

The CELEBS (controlled electrodynamic levitation and extraction of bioaerosols onto a substrate) platform addresses several limitations by enabling the examination of pathogen viability over short timescales, ranging from a few seconds to several minutes.^22,23^ Using this droplet levitation system, researchers observed a near-instantaneous decline in infectivity in 50% of SARS-CoV-2-laden droplets exposed to low RH.^23^ Despite its strengths, the levitation platform has a significant limitation. Biological droplets are inherently polydisperse, ranging from tens of nanometres to hundreds of micrometres.^24^ Moreover, pathogen decay rates within droplets may vary with size and volume; however, single-sized droplet levitation systems cannot capture the effects of variable droplet sizes on pathogen viability.^25^ Furthermore, levitated droplets equilibrate to sizes 2-4 times larger than those reported to contain over 90% of the detected SARS-CoV-2 RNA.^26,27^ Thus, highlighting the need for studies to assess pathogen viability within physiologically representative size ranges.

To meet these needs, we developed an aerosol platform to size-fraction and collect viable pathogen-laden droplets (VPLD) shortly after equilibration to indoor conditions (**Figure 1**). We used Phi6, a bacteriophage surrogate for SARS-CoV-2, to detect VPLD after equilibration.^28^ Furthermore, we selected Phi6 concentrations of 1×10^5^ PFU/mL and 5×10^5^ PFU/mL to reflect sputum RNA concentrations in infectious SARS-CoV-2 patients.^29^ Phi6 suspensions were prepared in 1.5, 3, or 6% weight-per-volume (w/v) peptone water solutions to simulate the non-volatile solute content in airway mucus lining in normal and moderate-to-severe muco-obstructive airway disease states.^17^ Atomisation of the suspensions generated six droplet fractions representing different sites of origin within the airways (initial diameter range, 1.73 to ≥20.28 µm).^30,31^ Trials were conducted during the temperate summer and winter to assess how seasonal variations in indoor RH impact pathogen viability.^16^ Using the gathered data, we discuss the potential effects of respiratory droplet size ranges, airway mucus composition, sputum viral loads, and seasonal variations in RH on indoor short-range transmission dynamics.

**Figure 1.**
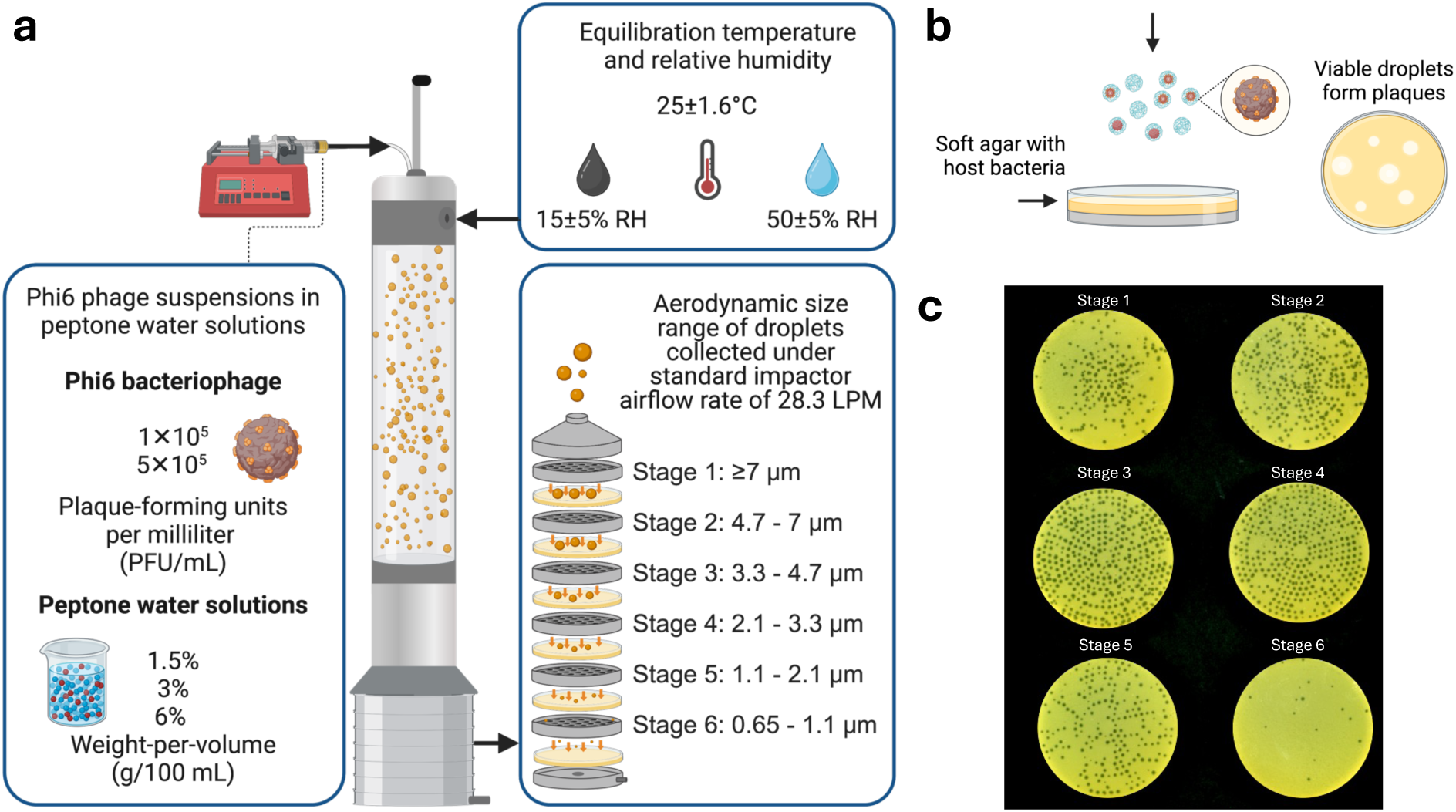
Schematic of the aerosol platform and the experimental workflow. (**a**) The platform contains three main components: the Blaustein single-jet atomizer, a 60-cm-high aerosol mixing chamber, and a 6-stage viable Andersen cascade impactor. Phi6 concentrations (1×10^5^ and 5×10^5^ plaque-forming units per millilitre (PFU/mL)) reflect sputum RNA concentrations in SARS-CoV-2 patients. Peptone water concentrations reflect the solute content of the airway mucus content. Low (15±5%) and intermediate (50±5%) relative humidity at 24±1.6°C reflects seasonal fluctuations in indoor environments. (**b**) The schematic depicts the viable pathogen-laden droplet detection method. The deposition of Phi6 on soft agar containing the host bacteria *Pseudomonas syringae* results in plaque formation owing to lytic phage activity. (**c**) Scanned image of six collection plates with plaques at the site of impaction of infective Phi6 aerosol droplets. Plaque counts were converted to coincidence-corrected whole number values to determine the recovered counts of viable pathogen-laden droplets. Schematics created with Biorender.

## 5. MATERIALS AND METHODS

### 5.1. Solutions

Tryptic soy broth (TSB) contained 30 g/L of tryptic soy broth (BD B211825). Tryptic soy agar (TSA) contained 30 g/L of tryptic soy broth (BD B211825) and 15 g/L of agar (Fisher BioReagents BP1423-500). Tryptic soy broth with yeast extract and magnesium sulphate (TSBY+) contained 30 g/L of tryptic soy broth (BD B211825), 6 g/L of yeast extract (Amresco J850-500G), and 0.6 g/L of magnesium sulphate (Amresco E797-500). Tryptic soy agar with yeast extract and magnesium sulphate (TSAY+) contained 30 g/L of tryptic soy broth (BD B211825), 6 g/L of yeast extract (Amresco J850-500G), 6 g/L of agar (Fisher BioReagents BP1423500), and 0.6 g/L of magnesium sulphate (Amresco E797-500). Peptone water solutions contained 15 g/L, 30 g/L, and 60 g/L of peptone water (Thermo Scientific CM0009) in ultrapure water. All solutions were sterilised by autoclaving (Yamato Steam Steriliser SM301) at 121°C for 20 minutes.

### 5.2. Bacteria and bacteriophage

*Pseudomonas* bacteriophage Phi6 (DSM 21518) is an accepted surrogate for SARS-CoV-2.^28^ The host bacterial strain, *Pseudomonas syringae* (DSM 21482), was cultured according to previously established methods with minor modifications.^28^ *P. syringae* was grown in TSBY media in a shaking incubator (25°C, 200 rpm, 48 hours). The 48-hour culture of *P. syringae* (80 µL) was diluted in fresh TSBY broth (4 mL) and incubated under agitation to prepare a subculture (25°C, 200 rpm, 6 hours). A single Phi6 plaque isolate was introduced into the subculture and incubated again (25°C, 200 rpm, 14 hours). The phage suspension was centrifuged (7000×*g*, 20 minutes, 4°C), and the supernatant (containing Phi6 bacteriophage) was passed through a 0.22-µm-pore-sized filter and stored at 4°C.

### 5.3. Preparation of bacteriophage feed solution

1×10^5^ PFU/mL and 5×10^5^ PFU/mL Phi6 suspensions in peptone water solutions (1.5%, 3%, or 6% w/v) used the phage stock suspension (3×10^10^ PFU/mL). The standard dilution plating method confirmed the concentration of Phi6 in peptone water suspensions.^32^ Briefly, the phage suspension was serially diluted in 900 µL aliquots of TSBY broth. An appropriate dilution was plated (100 µL) on bacterial agar plates in triplicate. The plates incubated at 25°C for 24 hours produced countable plaques (used to verify the titer of the suspension).

### 5.4. Phage aerosol detection and quantification

Bacterial agar plates (Pyrex 3160-100), prepared with 20 mL of TSA medium in the bottom layer and 7 mL of *P. syringae* overnight culture (15 mL) mixed with liquid TSAY media (100 mL) on top (27 mL total volume), were used to collect and quantify recovered counts of VPLD.^26^ Plates positioned below each stage collected size-fractioned aerosol droplets within the impactor and incubated at 22±2°C. Droplets with sufficient momentum travel through the stage perforation and impinge on the bacterial lawn plate positioned below the stage. Viable pathogen-laden droplets produced plaques after 24 to 36 hours. Plaque counts corresponding to each impactor stage were corrected for coincidence errors and rounded to the nearest whole number.^33,34^ For additional information, please refer to **Supporting Information Tables S19-S54**.

### 5.5. Fluorescein deposition trial

The deposition trials used phage-free peptone water solutions (1.5%, 3%, and 6% w/v) containing 1 mg/mL fluorescein sodium salt (Sigma-Aldrich F6377-100G). The solutions were atomised for 1 minute using 1.5 L/min of compressed airflow. Foil substrates were placed on 27 mL agar-containing plates to collect the droplets. The deposited material was washed from the foils using 5 mL of ultrapure water. Spectrophotometric measurements of fluorescein in 200 µL samples, added in triplicate to 96-well microplates (Greiner Bio-One 655086), used excitation and emission wavelengths of 490±5 and 515±5 nm (Biotek Synergy Neo2). For additional information, please refer to **Supporting Information Tables S7-S18.**

### 5.6. Statistical analysis and data visualisation

Nominal doses of fluorescein collected below individual impactor stages were analysed using one-way ANOVA, followed by Tukey’s multiple comparisons test (**Tables S13-S18**). The recovered counts of VPLD were analysed using a parametric statistical approach using two-way ANOVA, followed by Tukey’s multiple comparisons tests defined as * - *p*<0.05, ** - *p*<0.01, *** - *p*<0.001, **** - *p*<0.0001 (**Tables S41-S54**). The two-way ANOVA, a parametric statistical test, was selected because all 76 replicate datasets passed the Shapiro-Wilk and Kolmogorov-Smirnov normality tests (**Figure S4**). Based on the results of these normality tests, we cannot reject the null hypothesis that the 76 datasets do not follow a normal or Gaussian distribution. For additional information, please refer to **Supporting Information Figures S4-S6**. GraphPad 10 was used to perform all statistical analyses.

## 6. THEORY AND MODEL DEVELOPMENT

### 6.1. Phage aerosol generation and collection

The generation, equilibration, and collection of size-fractioned, viable phage-laden droplets (VPLD) occurred within the aerosol platform (**Figure 1**). The single-jet Blaustein generated polydisperse aerosol droplets from 0.17 mL of phage suspension for one minute. The droplets equilibrate for less than 20 seconds to ambient indoor RH within a 60-cm-high by 8-cm-wide glass aerosol mixing chamber. Airflow velocities, ranging from 0.05 to 0.18 m/s, transported the droplets down the length of the glass aerosol mixing chamber. For comparison, the time-averaged airflow velocity when speaking was reported in one study to be 0.208 m/s.^35^ A viable six-stage Andersen cascade impactor, positioned at the base of the aerosol mixing chamber, size-fractionates the droplets. The recovered counts of VPLD are determined using plaque-forming units on collection plates after incubation. For additional information, please refer to **Supporting Information Section S1, Tables S1-S6.**

### 6.2. Initial diameter range under the standard impactor airflow rate

The equilibrated size of droplets depends on the initial droplet size, RH, and solute content. The initial diameter of droplets, d_o_ in µm, is calculated according to **Equation 1**: d_50_ is the equilibrated droplet diameter in µm, C is the solute concentration in g⁄L, ρ is the density of water in g⁄L, and RH is the relative humidity.^36,37,38,39^ The d_50_ value is the reference cut-off range for the impactor stage. When operated at a standard volumetric airflow rate of 28.3 LPM, the impactor collects droplets within the following aerodynamic size fractions: 0.65-1.1, 1.1-2.1, 2.1-3.3, 3.3-4.7, 4.7-7, and ≥7 µm.

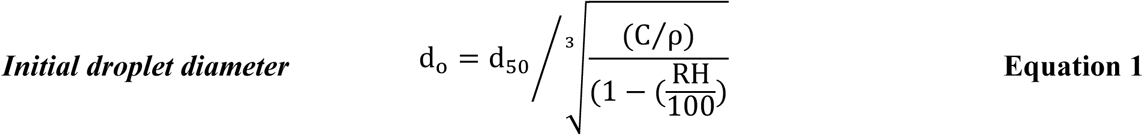

As illustrated by **Equation 1** and **Figure S1**, the initial diameter (d_o_) of droplets differs depending on the stage cut-off value (d_50_), the solute content (1.5, 3, or 6% w/v peptone water concentration), and equilibration condition (24±1.6°C; 15±5% or 50±5% RH) within the 60-cm-high aerosol mixing chamber. For example, droplets collected on the second stage of the impactor with a 4.7 µm cut-off diameter have an initial size of 18.05 µm if generated from 1.5% w/v peptone water solution and equilibrated at 15±5% RH or an initial size of 9.53 µm if generated from 6% w/v peptone water solution and equilibrated at 50±5% RH conditions (**Figure S1**). Adjusting the airflow rate of the Andersen impactor alters the cut-off diameters of the six stages, enabling the collection and quantification of VPLD with similar initial size distributions across 12 experimental conditions: three solute levels, two indoor RH settings, and two phage concentrations.

### 6.3. Initial diameter range under varied impactor airflow rates

We modified the impactor airflow rate, according to **Equation 2**, to collect equilibrated droplets with an initial diameter range of 1.73 to ≥20.28 µm across three solute levels and two RH conditions (**Figure S2, Tables S1-S6)**. This approach ensured the consistent collection of droplets of the same initial size range across impactor stages (under varied airflow rates) regardless of variations in RH conditions and solute levels (**Figure S1; Tables 1, 2**). Moreover, the rapid equilibration of droplets with initial sizes smaller than 50 µm, combined with the 3.4 to 12-second exposure period (depending on the impactor airflow rate), provided adequate time for droplets to equilibrate before collection.^12,38^

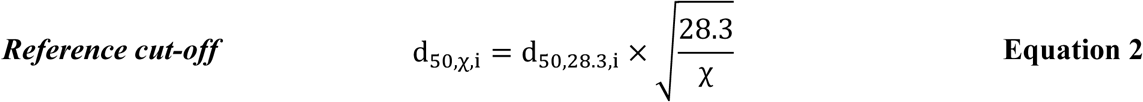

**Table 1.** Effect of varied impactor airflow rates on initial and equilibrated droplet sizes at 15±5% RH.

|  | **1.5% w/v** | **3% w/v** | **6% w/v** |
| --- | --- | --- | --- |
|  | **52.8 LPM** | **33 LPM** | **20.5 LPM** |
| **d<sub>o</sub> (μm)** | **d<sub>eq</sub> (μm)** |  |  |
| **≥18.65 to ≥20.28** | ≥5.12 | ≥6.48 | ≥8.22 |
| **12.52 to 20.28** | 3.44 to 5.12 | 4.35 to 6.48 | 5.52 to 8.22 |
| **8.79 to 13.62** | 2.24 to 3.44 | 3.06 to 4.35 | 3.88 to 5.52 |
| **5.59 to 9.56** | 1.54 to 2.24 | 1.94 to 3.06 | 2.47 to 3.88 |
| **2.93 to 6.09** | 0.81 to 1.54 | 1.02 to 1.94 | 1.29 to 2.47 |
| **1.73 to 3.19** | 0.48 to 0.81 | 0.6 to 1.02 | 0.76 to 1.29 |

**Table 2.** Effect of varied impactor airflow rates on initial and equilibrated droplet sizes at 50±5% RH.

|  | **1.5% w/v** | **3% w/v** | **6% w/v** |
| --- | --- | --- | --- |
|  | **38.5 LPM** | **24.2 LPM** | **15 LPM** |
| **d<sub>o</sub> (μm)** | **d<sub>eq</sub> (μm)** |  |  |
| **≥18.65 to ≥20.28** | ≥6 | ≥7.57 | ≥9.61 |
| **12.52 to 20.28** | 4.03 to 6 | 5.08 to 7.57 | 6.46 to 9.61 |
| **8.79 to 13.62** | 2.83 to 4.03 | 3.57 to 5.08 | 4.53 to 6.46 |
| **5.59 to 9.56** | 1.80 to 2.83 | 2.27 to 3.57 | 2.88 to 4.53 |
| **2.93 to 6.09** | 0.94 to 1.80 | 1.19 to 2.27 | 1.51 to 2.88 |
| **1.73 to 3.19** | 0.56 to 0.94 | 0.7 to 1.19 | 0.89 to 1.51 |

Equilibrated droplet size ranges were derived from the reference cut-off ranges for corresponding impactor stages.^40^ For a given stage, i, the cut-off diameter, d_50_, specifies the aerodynamic size of droplets collected with 50% efficiency.^40^ The impactor stage cut-off values are dependent on the airflow rate. Stage cut-off diameters for reference airflow rates, d_50,χ,i_, are calculated according to **Equation 2**, where χ is the reference airflow rate, i is the impactor stage, and d_50,28.3,i_, is the stage cut-off diameter under the standard airflow rate of 28.3 LPM.^40,41^ Reviews by Vuorinen et al. and Poydenot et al. indicate that evaporation time far exceeds settling time for droplets 20 µm and smaller, making this size range particularly relevant for understanding airborne transmission dynamics.^12,42^

### 6.4. Relevance and limitations of Phi6 as a surrogate for SARS-CoV-2

Phi6 is a biosafety level 1 organism that poses no risk of infection to humans and is frequently used as a surrogate for SARS-CoV-2 due to morphological and structural similarities. These include a phospholipid envelope, RNA genome, and comparable size (85 nm), as well as a spherical shape with surface projections.^28^ The use of Phi6 as a surrogate for SARS-CoV-2 to probe airborne transmission dynamics was based on a review of aerosol stability studies.

Parry-Nweye et al. reported that Phi6 within aerosols exhibited twofold higher decay rate at 75% RH compared to 45% RH, based on four viability measurements across 90 minutes.^43^ Similarly, Smither et al. observed that SARS-CoV-2 decayed at a rate of 0.91% per minute at 40-60% RH, which increased to 1.59% per minute at 68-88% RH at 19-22°C, using five viability measurements over 90 minutes.^44^ Moreover, Dabisch et al. reported a decay rate of 0.6% per minute while Schuit et al. reported a decay rate of 0.8% per minute for SARS-CoV-2-laden aerosols exposed to 20% RH at 20°C.^45,46^ This is consistent with the observations of Parry-Nweye et al. that the highest Phi6 viability within aerosols occurs at 25% RH.^43^

French et al. concluded Phi6 is less stable than SARS-CoV-2 at 40% RH.^25^ However, this was based on viability in 50 µL droplets (approximate diameter of 4.6 mm) (if spherical), in comparison to the 1.73 to ≥20.28 µm droplet diameters examined in this study.^25^ Additionally, viability was assessed after 4 and 8 hours of exposure, as compared to less than 20 seconds in the current work.^25^ Moreover, the concentration of Phi6 (10^6^ PFU/mL) and SARS-CoV-2 (10^5^ 50% tissue culture infective doses/mL) differed.^25^ Notably, the decay rate between Phi6 and SARS-CoV-2 was similar for the first hour of exposure.^25^ However, significant differences in decay rates emerged beyond two hours of exposure.^25^ French et al. reported no significant decay of Phi6 in 5 µL droplets exposed to 65% RH in the first 30 minutes.^25^ Similarly, Chin et al. reported that 5 µL droplets of SARS-CoV-2 did not significantly decay at 22°C and 65% RH during the first 30 minutes of exposure, based on four timepoints of sampling (1, 5, 10, and 30 minutes).^47^ These stability results indicate that Phi6 and SARS-CoV-2 exhibit comparable decay profiles over short exposure periods under similar environmental conditions.

In summary, Phi6 and SARS-CoV-2 exhibit similar stability profiles in aerosols in controlled temperature and humidity conditions, especially after short exposure timeframes. In contrast, virus stability data from large droplets taken over extended exposure periods with limited viability sampling produce divergent stability results between the two viruses. Experimental data from a surrogate organism, when considered alongside results from SARS-CoV-2 studies, epidemiological data, and clinical evidence, may provide novel insight into plausible mechanisms driving the short-range spread of infection.

## 7. RESULTS

### 7.1. Effect of RH, solute content, and impactor airflow rates on dose deposition profiles

To attribute differences in the counts of VPLD, collected under varying conditions such as RH, solute content, and impactor airflow rates, to changes in phage stability rather than delivery, we needed to ensure that these operating conditions did not affect the amount of material deposited below each stage. Consequently, the system was operated under each set of conditions using fluorescein-containing peptone water solutions. The fluorescein collected below each stage was quantified to determine the nominal dose delivered under different impactor airflow rates, RH conditions, and peptone water solutions.

Peptone water solutions containing 1 mg/mL of sodium fluorescein were aerosolised, and size-fractioned droplets were collected and quantified (**Tables S7-S12**). The results, presented in **Figure S3,** indicate nominal doses of aerosolised solution recovered from below each stage do not significantly vary despite variations in RH, solute content, and impactor airflow rates (**Tables S13-S18**). Supported by these findings, changes in phage viability within droplets, rather than variations in fractions of the aerosol reaching the collection plates, were used to interpret the results.

### 7.2. Recovered counts of VPLD collected below the first stage

**Figures 2a and 2b** present the VPLD collected below the first impactor stage, with an initial and equilibrated diameter range of ≥18.65 to ≥20.28 µm and ≥6 to ≥8.22 µm. The recovered VPLD, generated from 1×10^5^ PFU/mL phage suspensions, were not significantly affected by solute content or exposure to low versus intermediate RH (**Figure 2a; Tables S19-S29, S41**). However, recovered VPLD, generated from 5×10^5^ PFU/mL phage suspensions containing 6% w/v solute content and exposed to 50±5% RH, were significantly higher compared to the following: 1.5% w/v solute content and 15±5% RH; 6% w/v solute content and 15±5% RH; and 1.5% w/v solute content and 50±5% RH (**Figure 2b; Tables S30-S36, S47**).

**Figure 2.**
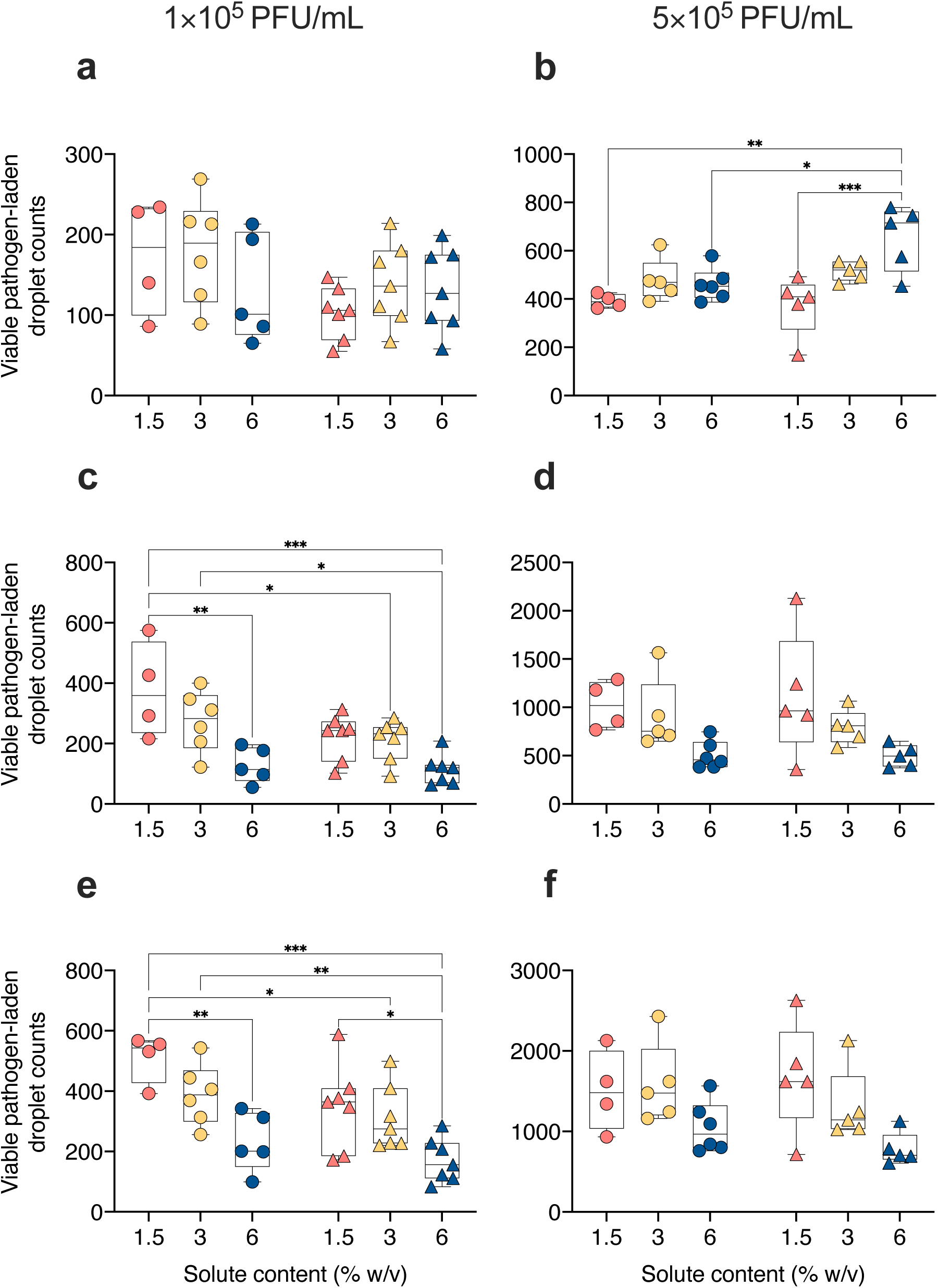
Effect of solute content, relative humidity, and phage concentration on recovered counts of viable pathogen-laden droplets collected below the first, second, and third impactor stages. Phi6 bacteriophage suspensions with concentrations of (**a,c,e**) 1×10^5^ PFU/mL and (**b,d,f**) 5×10^5^ PFU/mL were prepared in 1.5% (**coral**), 3% (**yellow**), and 6% (**blue**) w/v peptone water solutions and aerosolized into 15±5% (**circles**) and 50±5% (**triangles**) relative humidity conditions at 24±1.6°C. Graphs present coincidence-corrected counts of viable pathogen-laden droplets collected below the (**a,b**) first, (**c,d**) second, and (**e,f**) third impactor stages with initial size ranges of ≥18.65 to ≥20.28, 12.52 to 20.28, and 8.79 to 13.62 µm. Significance determined using two-way ANOVA followed by Tukey’s multiple comparisons tests defined as * - *p*<0.05, ** - *p*<0.01, *** - *p*<0.001.

### 7.3. Recovered counts of VPLD collected below the second stage

**Figures 2c and 2d** present the VPLD collected below the second impactor stage, with an initial and equilibrated diameter range of 12.52 to 20.28 µm and 4.03 to 8.22 µm. The recovered VPLD, generated from 1×10^5^ PFU/mL phage suspensions, containing 1.5% w/v solute content and exposed to 15±5% RH were significantly higher compared to the following: 6% w/v solute content and 15±5% RH; 3% w/v solute content and 50±5% RH; and 6% w/v solute content and 50±5% RH (**Figure 2c; Tables S19-S29, S42**). Additionally, recovered VPLD, generated from 1×10^5^ PFU/mL phage suspensions containing 3% w/v solute content and exposed to 15±5% RH, were significantly higher compared to droplets containing 6% w/v solute content and exposed to 50±5% RH (**Figure 2c; Tables S19-S29, S42**). Conversely, recovered VPLD generated from the 5×10^5^ PFU/mL phage suspensions were statistically comparable across the three solute levels and two exposure conditions (**Figure 2d; Tables S30-S36, S48**).

### 7.4. Recovered counts of VPLD collected below the third stage

**Figures 2e and 2f** present the VPLD collected below the third impactor stage, with an initial and equilibrated diameter range of 8.79 to 13.62 µm and 2.83 to 5.52 µm. The recovered VPLD, generated from 1×10^5^ PFU/mL phage suspensions containing 1.5% w/v solute content and exposed to 15±5% RH, were significantly higher compared to the following: 6% w/v solute content and 15±5% RH; 3% w/v solute content and 50±5% RH; and 6% w/v solute content and 50±5% RH (**Figure 2e; Tables S19-S29, S43**). Moreover, the recovered VPLD, generated from 1×10^5^ PFU/mL phage suspensions containing 3% w/v solute content and exposed to 15±5% RH, were significantly higher compared to recovered droplets containing 6% w/v solute content and exposed to 50±5% RH (**Figure 2e; Tables S19-S29, S43**). Lastly, recovered VPLD, generated from 1×10^5^ PFU/mL phage suspensions containing 1.5% w/v solute content and exposed to 50±5% RH, were significantly higher compared to recovered droplets containing 6% w/v solute content and exposed to 50±5% RH (**Figure 2e; Tables S19-S29, S43**). The recovered VPLD, generated from 5×10^5^ PFU/mL phage suspensions, were statistically comparable across the three solute levels and two exposure conditions (**Figure 2f; Tables S30-S36, S49**).

### 7.5. Recovered counts of VPLD collected below the fourth stage

**Figures 3a and 3b** present the VPLD collected below the fourth impactor stage, with an initial and equilibrated diameter range of 5.59 to 9.56 µm and 1.80 to 3.88 µm. The recovered VPLD, generated from 1×10^5^ PFU/mL phage suspensions containing 1.5% w/v solute content and exposed to 15±5% RH, were significantly higher compared to the following: 6% w/v solute content and 15±5% RH; 1.5% w/v solute content and 50±5% RH; 3% w/v solute content and 50±5% RH; and 6% w/v solute content and 50±5% RH (**Figure 3a; Tables S19-S29, S44**). Moreover, recovered VPLD, generated from 1×10^5^ PFU/mL phage suspensions containing 3% w/v solute content and exposed to 15±5% RH, were significantly higher compared to those containing 6% w/v solute content and exposed to 50±5% RH (**Figure 3a; Tables S19-S29, S44**). Conversely, the recovered VPLD generated from 5×10^5^ PFU/mL phage suspensions were similar across three solute levels and two exposure conditions (**Figure 3b; Tables S30-S36, S50**).

**Figure 3.**
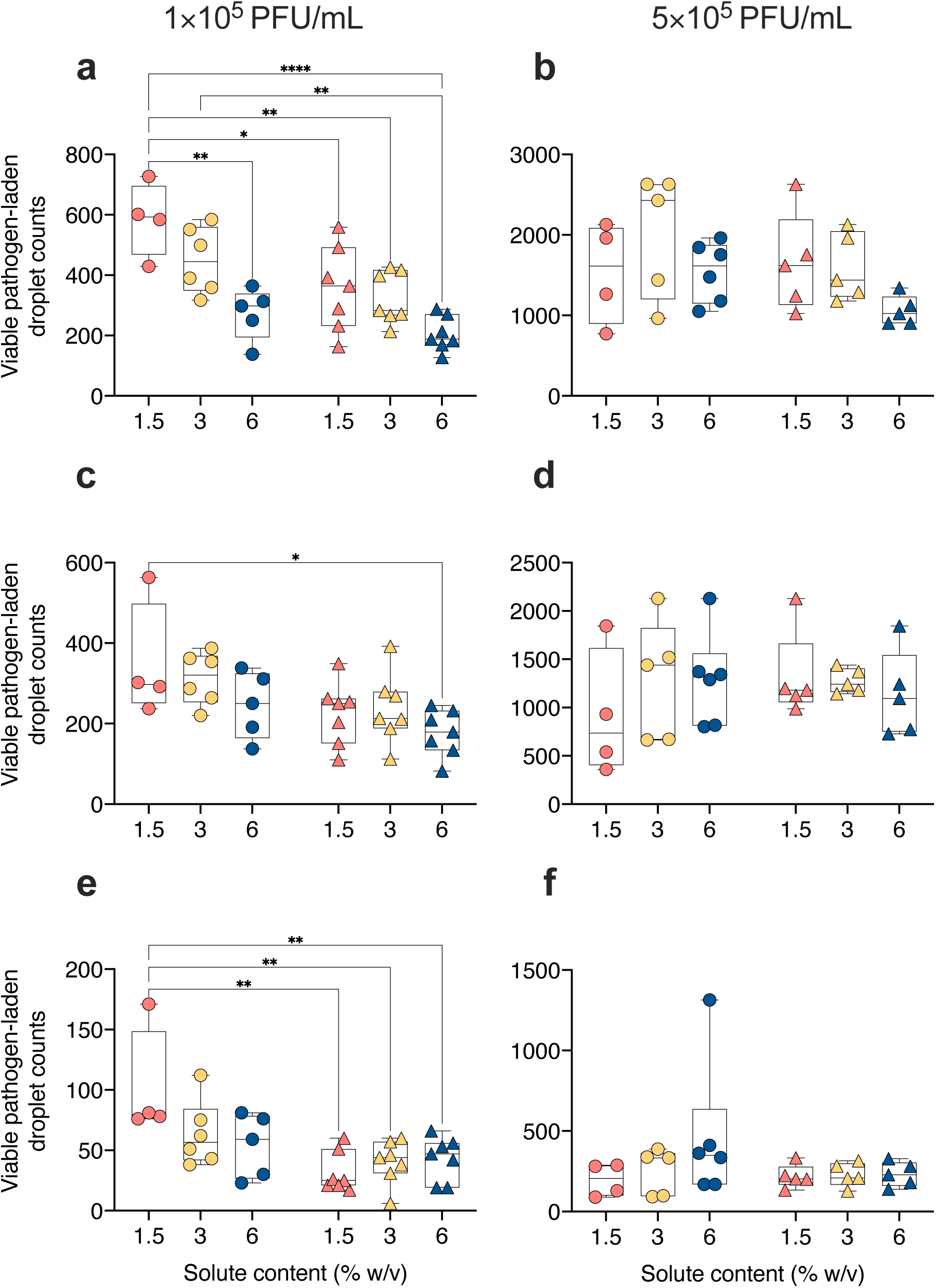
Effect of solute content, relative humidity, and phage concentration on recovered counts of viable pathogen-laden droplets collected below the fourth, fifth, and sixth impactor stages. Phi6 bacteriophage suspensions with concentrations of (**a,c,e**) 1×10^5^ PFU/mL and (**b,d,f**) 5×10^5^ PFU/mL were prepared in 1.5% (**coral**), 3% (**yellow**), and 6% (**blue**) w/v peptone water solutions and aerosolized into 15±5% (**circles**) and 50±5% (**triangles**) relative humidity conditions at 24±1.6°C. Graphs present coincidence-corrected counts of viable pathogen-laden droplets collected below the (**a,b**) fourth, (**c,d**) fifth, and (**e,f**) sixth impactor stage with initial size ranges of 5.59 to 9.56, 2.93 to 6.09, and 1.73 to 3.19 µm. Significance determined using two-way ANOVA followed by Tukey’s multiple comparisons tests defined as * - *p*<0.05, ** - *p*<0.01, **** - *p*<0.0001.

### 7.6. Recovered counts of VPLD collected below the fifth stage

**Figures 3c and 3d** present the VPLD collected below the fifth impactor stage, with an initial and equilibrated diameter range of 2.93 to 6.09 µm and 0.94 to 2.47 µm. The recovered VPLD, generated from 1×10^5^ PFU/mL phage suspensions containing 1.5% w/v solute content and exposed to 15±5% RH, were significantly higher compared to those containing 6% w/v solute content and exposed to 50±5% RH (**Figure 3c; Tables S19-S29, S45**). The recovered VPLD, generated from 5×10^5^ PFU/mL phage suspensions, were comparable across the solute levels and exposure conditions (**Figure 3d; Tables S30-S36, S51**).

### 7.7. Recovered counts of VPLD collected below the sixth stage

**Figures 3e and 3f** present the VPLD collected below the sixth impactor stage, with an initial and equilibrated diameter range of 1.73 to 3.19 µm and 0.56 to 1.29 µm. The recovered VPLD, generated from 1×10^5^ PFU/mL phage suspensions containing 1.5% w/v solute content and exposed to 15±5% RH, were significantly higher compared to the following: 1.5% w/v solute content and 50±5% RH; 3% w/v solute content and 50±5% RH; and 6% w/v solute content and 50±5% RH (**Figure 3e; Tables S19-S29, S46**). The recovered VPLD, generated from 5×10^5^ PFU/mL phage suspensions, were comparable across the solute levels and exposure conditions (**Figure 3f; Tables S30-S36, S52**).

## 8. DISCUSSION

### 8.1. Impact of solute concentration on viable pathogen-laden droplets

Epidemiological data indicate that 44% to 62% of SARS-CoV-2 transmission events occur during the presymptomatic phase, and peak during the early symptomatic phase, typically characterised by high viral loads.^48,49,50,51,52,53,54^ However, sputum production is rare during the early symptomatic phase of mild SARS-CoV-2 infection.^55^ During the progression of SARS-CoV-2 infection, mucus accumulates and viral loads decline.^56^ Importantly, respiratory secretions from ventilated SARS-CoV-2 patients revealed a significant elevation in solute levels similar to that observed in cystic fibrosis patients.^57,58^ To investigate the role of airway mucus composition and virus load on pathogen viability, VPLD generated from two different phage suspensions in three different solute concentrations was examined.^17^

Across three size fractions (initial diameter range, 5.59 to 20.28 µm), VPLD with low (1.5% w/v) solute content was significantly higher compared to those with a higher (6% w/v) solute content, after equilibrating to low winter humidity (15±5% RH) (**Figures 2c,2e,3a**). Additionally, in one size fraction (initial diameter range, 8.79 to 13.62 µm), VPLD with low solute content were significantly greater than those with high solute content (6% w/v) after equilibration to intermediate summer humidity (45±5% RH) **(Figure 2e)**. These results indicate that the pathogen viability is higher when embedded in a low-solute matrix shortly after equilibration. Similarly, decay of SARS-CoV-2 infectivity within aerosol droplets was higher when embedded in a matrix with a higher solids content.^45^ Elevated solute content is associated with increased viscosity of the airway mucus.^17,59^ This increase in viscosity makes it more challenging for droplets to form through shear-stress-induced breakup of the mucus lining, reducing the number of aerosol droplets.^20,60^ The pathophysiological changes associated with the late symptomatic phase of infection and severe COVID-19 may contribute to impaired droplet formation and decreased pathogen viability within aerosol droplets, to reduce the likelihood of airborne transmission in the later stages of severe disease.

### 8.2. Impact of increased sputum viral loads on viable pathogen-laden droplets

Interestingly, VPLD generated from concentrated phage suspensions (5×10^5^ PFU/mL), did not differ across the three solute levels and two RH conditions in solute concentrations, in five of the six fractions (initial diameter range, 1.73 to 20.28 µm) (**Figures 2d,2f,3b,3d,3f**). Similar recovery of VPLD across favourable (low solute/low RH) and unfavourable conditions (elevated solute/intermediate RH) suggests a protective effect of elevated viral loads on viability within aerosol droplets. The slower decay of Phi6 with increases in concentration was previously reported and may be due to concentration-dependent virus aggregation.^61,62^ Notably, a high viral load, characterising the early symptomatic phase of SARS-CoV-2 infection, may function as a buffer against high-solute- and intermediate-RH-dependent reduction in viability under less favourable conditions.

SARS-CoV-2, like many other respiratory viral infections, has become endemic and continues to evolve and persist with time.^64^ Clinical data indicates the evolved SARS-CoV-2 variant, Delta, produces higher peak sputum viral loads than ancestral strains.^50^ The higher viral load typical of the early symptomatic phase may counteract the effects of elevated solute levels on pathogen viability within aerosol droplets. However, in vaccinated individuals, peak viral loads during the early symptomatic phase are dampened.^54,63^ The lower viral loads in vaccinated individuals may limit the benefit of enhanced aerosol viability during the early symptomatic phase. Pathogen-load-dependent shifts in pathogen viability within droplets may alter transmissibility during the summer months (intermediate indoor humidity) or the infectiousness of individuals with muco-obstructive airway conditions (high solute levels in mucus).^16,17^ Understanding the viral loads produced by VOCs in vaccinated and unvaccinated individuals could help inform public health policies.

### 8.3. Impact of seasonal variations in indoor relative humidity on viable pathogen-laden droplets

SARS-CoV-2 infection rates typically peak during temperate winter and wane during temperate summer, characterised by low and intermediate indoor RH.^16,65^ The recovery of VPLD, equilibrated under indoor conditions during temperate summer and winter, was used to probe the effect of seasonal variations in indoor RH on pathogen viability within aerosol droplets. The recovered counts of VPLD (initial diameter range, 5.59 to 9.56 and 1.73 to 3.19 µm) containing low solute content (1.5% w/v) and equilibrated to indoor conditions during temperate winter (15±5% RH) were significantly higher than those equilibrated during temperate summer (50±5% RH) (**Figures 3a,3e**). Moreover, recovered counts of VPLD (initial diameter range, 1.73 to 20.28 µm) containing low solute content (1.5% w/v) and equilibrated to indoor conditions during temperate winter (15±5% RH) were significantly higher than high solute content (6% w/v) and equilibrated to indoor conditions during temperate summer (50±5% RH) (**Figures 2c,2e,3a,3c,3e**).

The contrast between the airway conditions (37°C and 99.5% RH) and the external environment drives the transformations of respiratory droplets outside the airways.^26,66^ Low indoor RH during temperate winters accelerates evaporation and rapidly concentrates non-volatile solutes such as salts and proteins.^23^ Under these conditions, salts become sequestered within crystalline structures, excluding the free diffusion of these ions, which may mitigate salt toxicity-induced viral inactivation.^23,67,68^ However, the salts in droplets exposed to intermediate RH (higher droplet-phase water activity) may contribute to increased viral inactivation in temperate summer months.^67,69^ Furthermore, equilibration and the formation of salt crystal inclusions could be delayed by high protein levels.^70^ Thus, equilibration under intermediate RH and high protein levels may delay salt crystallisation and increase pathogen inactivation to lower the risk of transmission during temperate summer. The described microphysical processes agree with our experimental findings of higher recovery of VPLD under low solute and low RH conditions compared to high solute and intermediate RH conditions.

The relative importance of host- and pathogen-related factors contributing to seasonal variations in respiratory virus infection rates is unresolved.^16^ Behavioural changes in winter, such as increased time spent indoors and reduced ventilation due to efforts to insulate against the cold, may lead to the accumulation of infectious aerosols and increase transmission risk.^16^ Additionally, the viability of pathogens within aerosol droplets exposed to indoor conditions during summer versus winter may also impact transmission risk.^16^

A recent animal study showed that increasing the ventilation rates, from an air exchange rate of 1.3 to 23 air changes per hour, failed to reduce transmission during frequent close-contact interactions.^71^ Further to this point, the close-contact scenario described by Zhang et al. illustrates why ventilation might be ineffective at reducing short-range transmission risk.^72^ During a face-to-face conversation at a distance of 1 m, assuming breathing and speaking velocities of 2.4 and 3.9 m/s, a susceptible individual may inhale droplets directly from the exhaled plume of an infectious talking subject within 0.4 seconds.^72^ Given how quickly an individual could inhale infectious droplets during short-range face-to-face interactions, indoor ventilation conditions in summer versus winter may not significantly influence exposure to infectious droplets. Based on this discussion and our findings, the concentration of VPLD present shortly after their release may play a role in short-range transmission dynamics and influence seasonal oscillations in the airborne spread of respiratory viruses. However, close-contact interactions are not the only means of SARS-CoV-2 spread; improved ventilation may significantly reduce the risk of long-range transmission events mediated by the accumulation of infectious droplets over time.^73^

### 8.4. Impact of respiratory droplet sizes on viable pathogen-laden droplets

The results for droplet fractions, with an initial droplet diameter range of 1.73 to 20.28 µm, are consistent with the discussed mechanisms. Specifically, salt crystallisation under low RH conditions may reduce solute toxicity, improving the recovery of VPLD generated from the low-phage suspension. Meanwhile, virus aggregation may function as a buffer against high solute and intermediate RH-dependent viability changes in droplets generated from the high-phage suspension.

However, VPLD with an initial diameter of ≥20.28 µm do not follow these trends (**Figures 2a and 2b)**. Solute levels and RH conditions did not significantly impact the recovery of VPLD generated from the low-phage suspensions (**Figure 2a**). Different trends emerged when analysing droplets generated from 5-fold concentrated phage suspensions, where the recovery of VPLD was significantly higher in the high solute/intermediate RH condition than in the low solute/low RH condition (**Figure 2b**). The recovery of VPLD containing a high solute content (6% w/v) was significantly higher compared to low solute content (1.5%) after equilibration to indoor conditions during temperate summer (50±5% RH) (**Figure 2b**). Haddrell et al. reported significantly greater stability of the SARS-COV-2 Delta variant of concern in a matrix of 3xMEM media as compared to 1xMEM media when exposed to 40% RH for less than 5 seconds.^74^ It is interesting to note that a similar matrix-viability relationship in Phi6-containing droplets with an equilibrated diameter of ≥8.22 µm, just below the 10 µm equilibrated droplet diameter investigated by Haddrell et al.^74^

The discrepant viability trends between the two size fractions may result from the rate of equilibration experienced by each size fraction.^75^ The formation of concentration gradients within equilibrating droplets is more common within larger droplets.^75^ These gradients result from phase separation, creating distinct microenvironments enriched in organic proteins or inorganic salts.^23,68,70,76^ During droplet equilibration, the slower diffusion of large protein molecules compared to ions produces a protein-enriched shell surrounding a salt-enriched core.^68,77^ The location of pathogens within these microenvironments may influence viability outcomes, with protein-enriched regions enhancing and salt-enriched regions reducing it.^23,68,78,79^

The non-preferential localisation of phage within protein- or salt-enriched microenvironments could account for the statistically similar recovery of VPLD from the low phage suspensions across all six conditions. Conversely, the preferential localisation of phage within protein-enriched microenvironments under high solute/intermediate RH conditions could account for the significantly higher recovery of VPLD generated from the high phage suspensions. Our findings indicate that viral aggregation, under increased phage concentrations, may favour localisation within protein-enriched microenvironments.

## 9. LIMITATIONS AND FUTURE DIRECTIONS

This study has limitations related to the experimental design and the instrumentation that warrant discussion. The bacteriophage Phi6, rather than SARS-CoV-2, was used in experimental trials. Phi6 shares morphological and structural similarities with SARS-CoV-2 and comparable environmental stabilities within aerosols over short periods.^25^ However, Phi6 data alone cannot provide meaningful insight into the transmission of COVID-19. Consequently, Phi6-based results on pathogen viability within aerosol droplets, in conjunction with epidemiological data, clinical evidence, and in vivo and in vitro studies using SARS-CoV-2, were used to describe plausible mechanisms driving short-range airborne transmission dynamics. To extend the relevance of the findings presented in this study, an attenuated SARS-CoV-2 strain could be used in place of Phi6, followed by droplet collection on an appropriate collection medium within the impactor.^80^ This approach may help clarify the value of Phi6-based data in understanding SARS-CoV-2.

The medium used to prepare the bacteriophage suspensions is a simplified substitute for the solute content in airway mucus. However, this medium lacks the complexity of respiratory secretions and may impact pathogens differently from peptone water solution.^31^ However, there is considerable variability in the composition of biological samples.^21^ While peptone water serves as a practical alternative for experimental purposes, it may not accurately reflect the transformations droplets derived from human respiratory secretions may undergo. Replicating this work with sputum samples could add to the clinical relevance of the findings from this study.

Impactor airflow rates were varied to collect roughly the same size fractions across solute levels and RH conditions. The fluorescein trial data indicate that the nominal dose amounts reaching each stage did not vary significantly with changing airflow rates. However, changes to the impactor airflow rate alter the impaction velocity, potentially affecting the viability and collection efficiency of the bacteriophage-laden droplets, especially in lower stages.^81^ A softer and more viscous impaction surface was created by lowering the agar content of the top layer of the collection medium (the bacterial lawn) to minimise droplet bounce and reduce the stress of impaction across varied airflow rates. Furthermore, the two-minute trial period limits the evaporation and desiccation of the collection medium.

The initial droplet size ranges were estimated using an established formula and input values that included impactor cut-off values (calculated for non-standard impactor airflow rates), solute content, and ambient RH and temperature.^36,37,38,39^ Mathematical approaches that account for airflow velocity, thermodynamic changes, and solute distribution within droplets when calculating the initial size of aerosol droplets are available but were beyond the scope of this work.^82,83^

The aerosol platform used in this study for data collection has limitations. Only 10-20% of the aerosol droplets reach the impactor, with substantial losses within the glass aerosol mixing chamber and the impactor (excluding the surface of the collection plates). Additionally, our experimental setup is limited to endpoint collection of size-fractionated, viable pathogen-laden droplets. The volumetric flow rate and the height of the aerosol mixing chamber dictate how long aerosols exist within the platform. As such, the platform cannot maintain aerosols for specific time intervals to assess viability over time. To enable continuous or time-resolved monitoring of viral decay in aerosols, future iterations of this platform could incorporate a Goldberg rotating drum or an electrodynamic balance integrated with the aerosol chamber and impactor. These modifications would allow for prolonged aerosol suspension under controlled conditions and facilitate kinetic studies of viral viability. While the design and validation of such an expanded platform are beyond the scope of the present study, this is a promising avenue for future research.

SARS-CoV-2 RNA concentrations in sputum can reach 1×10^11^ PFU/mL.^29^ In this study, droplets were generated for one minute from phage suspensions at concentrations of 1×10^5^ and 5×10^5^ PFU/mL, dispensed at a feed flow rate of 170 µL/min, using compressed air delivered at an airflow rate of 1.5 L/min. These operating parameters resulted in the recovery of countable droplets on collection plates within the impactor. However, to quantify VPLD generated from more concentrated suspensions, operating parameter values must be adjusted to prevent over-saturation of the collection plates, which hinders reliable quantification.^84^ Researchers interested in droplets generated from concentrated suspensions should consider the following adjustments to operating conditions: a shorter atomisation period, reduced feed flow rates, and reduced airflow rates. These modifications may allow the collection of countable droplets generated from concentrated feed suspension.

Lastly, this study focused on droplets with an initial diameter ranging from 1.73 to ≥20.28 µm, as these size fractions equilibrate much faster than they settle.^12^ However, initial droplet sizes as large as 80 µm, falling from a height of 1.5 meters, may equilibrate over various indoor conditions to remain airborne indefinitely.^85^ Thus, future studies examining pathogen viability in droplet fractions between 20 and 80 µm could provide a more comprehensive picture of short-range airborne transmission dynamics.

## 10. CONCLUSION

This study evaluated the effects of four key factors – respiratory droplet size, mucus composition, viral load, and indoor relative humidity – on the recovery of viable pathogen-laden droplets (VPLD). The novelty of this work lies in the examination of pathogen viability within droplets, with an initial diameter range of 1.73 to ≥20.28 µm, shortly after exposure (<20 seconds), to probe short-range transmission dynamics indoors, using a custom-designed impactor-based aerosol platform. Experimental findings obtained using the bacteriophage Phi6, in conjunction with epidemiological data, clinical evidence, and experimental studies using SARS-CoV-2, were used to describe potential mechanisms of short-range transmission.

Modelling low solute levels in the airway mucus lining, characteristic of the presymptomatic phase, increased recovery of VPLD (initial diameter range, 5.59 to 20.28 µm) in temperate winter (**Figures 2c,2e,3a**). The combination of low solute content in the airway lining and low RH in temperate winter may favour pathogen persistence within aerosol droplets, potentially elevating short-range transmission risk during the presymptomatic phase. This mechanism aligns with epidemiological data indicating higher transmission risk from asymptomatic, presymptomatic, and mildly symptomatic individuals before the onset of severe symptoms.^10^

Conversely, higher solute content, characteristic of the late symptomatic phase, was associated with reduced recovery of VPLD. Epidemiological data show a decline in transmission risk during later stages of SARS-CoV-2 infection.^10^ Moreover, high viral loads, characteristic of the early symptomatic phase, led to the comparable recovery of VPLD (initial diameter range, 1.73 to 20.28 µm) across all solute levels and RH conditions (**Figures 2d,2f,3b,3d,3f**). A higher viral load may mitigate the effects of high solute levels and intermediate RH to increase pathogen viability under less favourable conditions.

Pathogen viability within droplets, across seasonal variations in indoor RH, was also explored. The counts of VPLD (initial diameter range, 1.73 to 20.28 µm) with a low solute content and equilibrated to low RH (temperate winter) was significantly higher than those containing a high solute content and equilibrated to intermediate RH (temperate summer) (**Figures 2c,2e,3a,3c,3e**). Low RH accelerate droplet evaporation, promoting the sequestration of salts as crystal inclusions within droplets.^70^ These microphysical processes may enhance viability within aerosol droplets. In contrast, solutes remain dissolved under intermediate RH conditions typical of temperate summer, leading to salt-toxicity-induced viral inactivation. Our findings and proposed mechanisms align with seasonal variation in SARS-CoV-2 transmission rates.^16^

Lastly, our results suggest that respiratory droplet size influences the number of VPLD present shortly after expulsion. Larger droplets from the upper respiratory tract are more likely to form concentration gradients during the equilibration process, creating distinct microenvironments that may impact pathogen viability.^75^ Depending on the localisation of pathogens within protein-enriched or salt-enriched regions, it may enhance or compromise viability.^75^ In the low pathogen concentration, representing the presymptomatic phase, the collection of VPLD within the ≥20.28 µm size fraction was consistent across all solute levels and humidity conditions, suggesting non-specific localisation of virions within either protein- or salt-enriched compartment (**Figure 2a**). Under the high pathogen load condition, representing the early symptomatic phase, higher counts of VPLD in the high solute/intermediate RH condition indicate preferential localisation within protein-enriched compartments (**Figure 2b**). Virus aggregation behaviour within large equilibrating droplets may influence localisation within salt- or protein-rich microenvironments, possibly altering pathogen viability.

Together, these findings highlight the importance of ventilation and humidity control in indoor settings. Maintaining adequate ventilation and avoiding overly dry environments—especially during winter—may reduce the viability of airborne viruses. Early detection and isolation of presymptomatic or mildly symptomatic individuals, who may release more viable pathogens, remain critical. These strategies, informed by a mechanistic understanding of droplet physics and biology, may support more effective interventions.

## Data Availability

All data produced in the present study are available upon reasonable request to the authors

## 11. SUPPORTING INFORMATION

The following data sets are available as supporting information.

- Section S1. Platform operation
- Sections S2, S3. Spectrophotometric detection of doses of 1.5, 3, and 6% w/v peptone water solutions containing 1 mg/mL fluorescein, aerosolized and collected below each impactor stage under 15±5% and 50±5% RH conditions
- Section S4. One-way ANOVA followed by Tukey’s multiple comparisons test results for doses of 1.5, 3, and 6% w/v peptone water solutions containing 1 mg/mL fluorescein aerosolized and collected below each impactor stage
- Sections S5, S6. Recovered counts of viable pathogen-laden droplet counts generated from 1×10^5^ PFU/mL phage suspensions collected under 15±5% and 50±5% RH
- Sections S7, S8. Recovered counts of viable pathogen-laden droplet counts generated from 5×10^5^ PFU/mL phage suspensions collected under 15±5% and 50±5% RH
- Sections S9. Normality test results for recovered counts of viable pathogen-laden droplets
- Section S10. Two-way ANOVA followed by Tukey’s multiple comparison test results for the recovered counts of viable pathogen-laden droplets collected below each impactor stage
- Section S11. Two-way ANOVA followed by Tukey’s multiple comparison test results for the recovered counts of viable pathogen-laden droplets collected below all impactor stages

## 12. ACKNOWLEDGEMENTS

This research was undertaken, in part, thanks to funding from the CIHR project grants program (Z.H., M.D.), Canada Research Chairs Program (Z.H.), and Natural Sciences and Engineering Research Council of Canada (NSERC) Discovery Grants Program (Z.H.). M.T. is funded partially by the Ontario Graduate Fellowship (OGF) Program through the Government of Ontario. Paul Gatt, Justin Bernar, John Colenbrander from the Chemical and Mechanical Engineering Machine Shop and Doug Keller were instrumental in helping to design and build the aerosol platform.

## 13. TABLE OF CONTENTS GRAPHIC

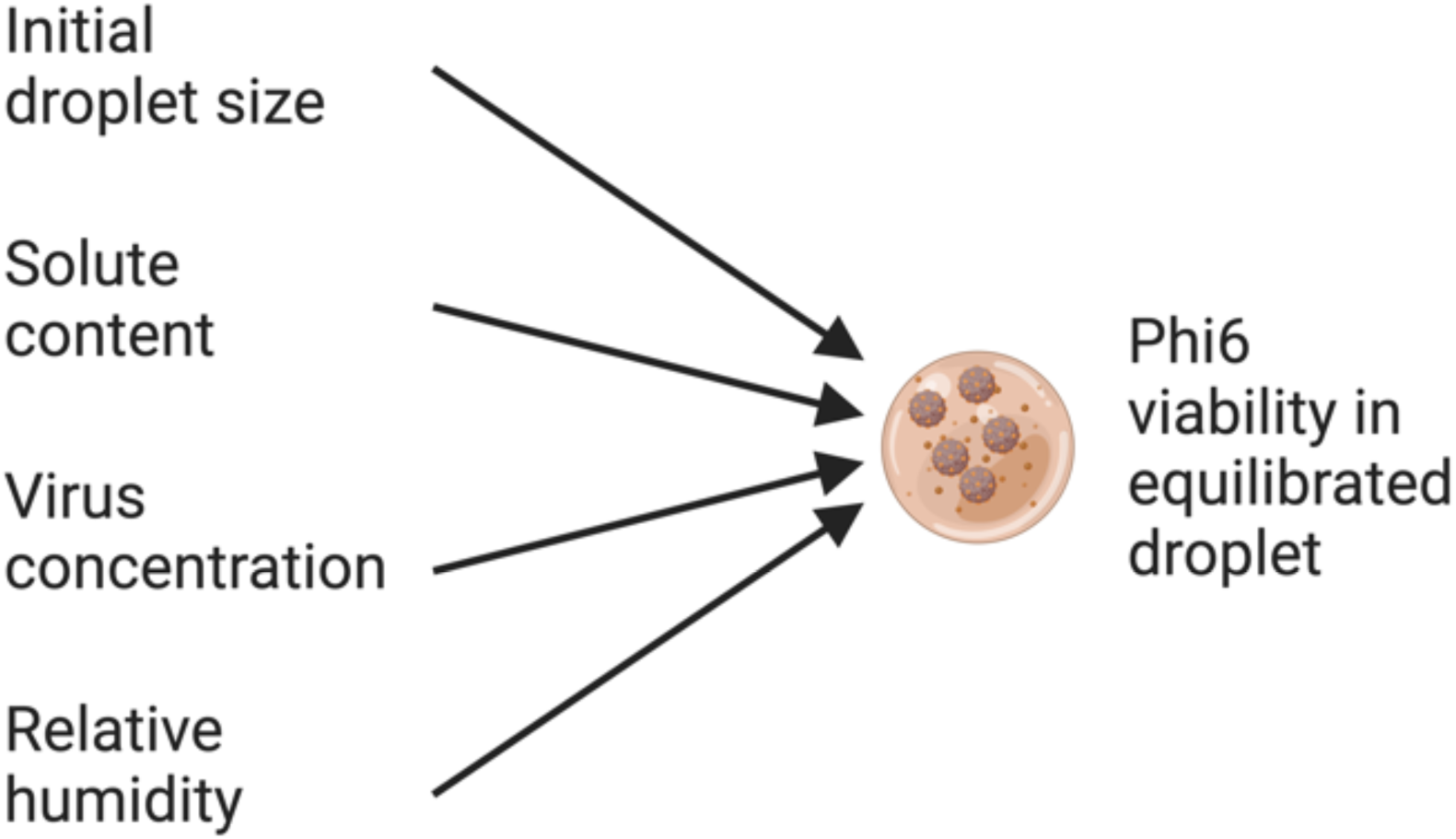

