## Supplementary Information for "An impactor-based aerosol platform for probing indoor, short-range transmission dynamics: a Phi6 bacteriophage study"

---

**TABLE OF CONTENTS**

Section S1. Platform operation

Section S2. Spectrophotometric detection of doses of 1.5, 3, and 6% w/v peptone water solutions containing 1 mg/mL fluorescein, aerosolized and collected below each impactor stage under 15±5 RH conditions

Section S3. Spectrophotometric detection of doses of 1.5, 3, and 6% w/v peptone water solutions containing 1 mg/mL fluorescein, aerosolized and collected below each impactor stage under 50±5% RH conditions

Section S4. One-way ANOVA followed by Tukey's multiple comparisons test results for doses of 1.5, 3, and 6% w/v peptone water solutions containing 1 mg/mL fluorescein aerosolized and collected below each impactor stage

Section S5. Recovered counts of viable pathogen-laden droplet counts generated from $1\times10^5$ PFU/mL phage suspension collected under 15±5% RH

Section S6. Recovered counts of viable pathogen-laden droplet counts generated from $1\times10^5$ PFU/mL phage suspension collected under 50±5% RH

Section S7. Recovered counts of viable pathogen-laden droplet counts generated from $5\times10^5$ PFU/mL phage suspension collected under 15±5% RH

Section S8. Recovered counts of viable pathogen-laden droplet counts generated from $5\times10^5$ PFU/mL phage suspension collected under 50±5% RH

Section S9. Normality test results for recovered counts of viable pathogen-laden droplets

Section S10. Two-way ANOVA followed by Tukey's multiple comparisons test results for recovered counts of viable pathogen-laden droplets collected below each impactor stage

Section S11. Two-way ANOVA followed by Tukey's multiple comparison test results for the recovered counts of viable pathogen-laden droplets collected below all impactor stages

**Number of pages: 50**
**Number of tables: 54**
**Number of figures: 6**

### SECTION S1. PLATFORM OPERATION

The main components of the aerosol platform consisted of a single-jet atomiser (CH Technologies ARGBLM2), a 60-cm-high by 8-cm-wide glass aerosol mixing chamber, and a six-stage viable Andersen cascade impactor (Tisch Environmental TE-10-800). [1,2] The Blaustein atomiser fitted with a 10-40 expansion plate (CH Technologies ARGBLM0031) generated polydisperse phage aerosols for one minute.[1] Phage suspension was dispensed from a 5 mL syringe (BD 309646) and delivered to the atomiser through 1/16'' x 3/16'' ID/OD silicone tubing (McMaster-Carr 51135K608). The liquid feed flow rate of 170 μL/min was maintained using a syringe pump (Fisherbrand 780100I). The compressed airflow rate of 1.5 L/min was maintained using a pressure gauge (IMI NORGREN R72G-2AT-RFG). Filtered room air entered the system at volumetric airflow rates of 13.5 to 51.3 L/min. The volumetric airflow rate of 15 to 52.8 L/min was maintained within the system using a vacuum pump (GAST 0823-101Q-G608NEX). Monitoring upstream and downstream airflow rates utilised OMEGA (FMA-A2317) and TSI (5300 series) flow meters, respectively. The ASTM International standard F2101-19 guided the initial design and operation of the platform.[3] A SensorPush device (HT. w 16859565) measured the RH and temperature within the platform before the start of each trial. The trials took place on days when RH was 50±5% (summer) or 15±5% (winter) at 24±1.6°C. Droplets experience flight times of <20 seconds within the chamber, under varied impactor airflow rates of 15 to 52.8 L/min, before reaching the impactor, followed by size-fractioning and collection. Sterile glass Petri dishes (Pyrex 3160-100) prepared with 20 mL of TSA medium in the bottom layer and 7 mL of *P. syringae* overnight culture (15 mL) mixed with liquid TSAY media (100 mL) on top (27 mL total volume) were used to collect and quantify recovered counts of VPLD.[26]

**Table S1. Equilibrated and initial droplet size calculation for 1.5% w/v peptone water suspensions collected under 15±5% RH at 52.8 LPM impactor airflow rate**

| Stage | $d_{50}$ 52.8 LPM | $d_o$ 10% RH | $d_o$ 15% RH | $d_o$ 20% RH | $\frac{d_{eq}}{d_{o,15\%}}$ |
|---|---|---|---|---|---|
| 1 | ≥5.12 | ≥20.06 | ≥19.68 | ≥19.29 | 0.26 |
| 2 | 3.44 | 13.47 | 13.22 | 12.95 | 0.26 |
| 3 | 2.42 | 9.46 | 9.28 | 9.09 | 0.26 |
| 4 | 1.54 | 6.02 | 5.91 | 5.79 | 0.26 |
| 5 | 0.81 | 3.15 | 3.09 | 3.03 | 0.26 |
| 6 | 0.48 | 1.86 | 1.83 | 1.79 | 0.26 |

$d_{50,28.3,i}$ = 7, 4.7, 3.3, 2.1, 1.1, 0.65 for stages 1 to 6, respectively.
$d_{50}$ = Cut-off diameter; $d_{eq}$ = Equilibrated droplet diameter; $d_o$ = Initial droplet diameter

**Table S2. Equilibrated and initial droplet size calculation for 3% w/v peptone water suspensions collected under 15±5% RH at 33 LPM impactor airflow rate**

| Stage | $d_{50}$ 33 LPM | $d_o$ 10% RH | $d_o$ 15% RH | $d_o$ 20% RH | $\frac{d_{50}}{d_{o,15\%}}$ |
|---|---|---|---|---|---|
| 1 | ≥6.48 | ≥20.14 | ≥19.76 | ≥19.37 | 0.33 |
| 2 | 4.35 | 13.52 | 13.27 | 13.00 | 0.33 |
| 3 | 3.06 | 9.50 | 9.32 | 9.13 | 0.33 |
| 4 | 1.94 | 6.04 | 5.93 | 5.81 | 0.33 |
| 5 | 1.02 | 3.17 | 3.11 | 3.04 | 0.33 |
| 6 | 0.60 | 1.87 | 1.84 | 1.80 | 0.33 |

**Table S3. Equilibrated and initial droplet size calculation for 6% w/v peptone water suspensions collected under 15±5% RH at 20.5 LPM impactor airflow rate**

| Stage | $d_{50}$ 20.5 LPM | $d_o$ 10% RH | $d_o$ 15% RH | $d_o$ 20% RH | $\frac{d_{50}}{d_{o,15\%}}$ |
|---|---|---|---|---|---|
| 1 | ≥8.22 | ≥20.28 | ≥19.90 | ≥19.50 | 0.41 |
| 2 | 5.52 | 13.62 | 13.36 | 13.09 | 0.41 |
| 3 | 3.88 | 9.56 | 9.38 | 9.19 | 0.41 |
| 4 | 2.47 | 6.09 | 5.97 | 5.85 | 0.41 |
| 5 | 1.29 | 3.19 | 3.13 | 3.06 | 0.41 |
| 6 | 0.76 | 1.88 | 1.85 | 1.81 | 0.41 |

**Table S4. Equilibrated and initial droplet size calculation for 1.5% w/v peptone water suspensions collected under 50±5% RH at 38.5 LPM impactor airflow rate**

| Stage | $d_{50}$ 38.5 LPM | $d_o$ 45% RH | $d_o$ 50% RH | $d_o$ 55% RH | $\frac{d_{50}}{d_{o,50\%}}$ |
|---|---|---|---|---|---|
| 1 | ≥6.00 | ≥19.94 | ≥19.31 | ≥18.65 | 0.31 |
| 2 | 4.03 | 13.39 | 12.97 | 12.52 | 0.31 |
| 3 | 2.83 | 9.40 | 9.11 | 8.79 | 0.31 |
| 4 | 1.80 | 5.98 | 5.79 | 5.59 | 0.31 |
| 5 | 0.94 | 3.13 | 3.04 | 2.93 | 0.31 |
| 6 | 0.56 | 1.85 | 1.79 | 1.73 | 0.31 |

**Table S5. Equilibrated and initial droplet size calculation for 3% w/v peptone water suspensions collected under 50±5% RH at 24.2 LPM impactor airflow rate**

| Stage | $d_{50}$ 24.2 LPM | $d_o$ 45% RH | $d_o$ 50% RH | $d_o$ 55% RH | $\frac{d_{50}}{d_{o,50\%}}$ |
|---|---|---|---|---|---|
| 1 | ≥7.57 | ≥19.96 | ≥19.34 | ≥18.67 | 0.39 |
| 2 | 5.08 | 13.40 | 12.98 | 12.53 | 0.39 |
| 3 | 3.57 | 9.41 | 9.12 | 8.80 | 0.39 |
| 4 | 2.27 | 5.99 | 5.80 | 5.60 | 0.39 |
| 5 | 1.19 | 3.14 | 3.04 | 2.93 | 0.39 |
| 6 | 0.70 | 1.85 | 1.80 | 1.73 | 0.39 |

**Table S6. Equilibrated and initial droplet size calculation for 6% w/v peptone water suspensions collected under 50±5% RH at 15 LPM impactor airflow rate**

| Stage | $d_{50}$ 15 LPM | $d_o$ 45% RH | $d_o$ 50% RH | $d_o$ 55% RH | $\frac{d_{50}}{d_{o,50\%}}$ |
|---|---|---|---|---|---|
| 1 | ≥9.61 | ≥20.12 | ≥19.49 | ≥18.82 | 0.49 |
| 2 | 6.46 | 13.51 | 13.09 | 12.64 | 0.49 |
| 3 | 4.53 | 9.49 | 9.19 | 8.87 | 0.49 |
| 4 | 2.88 | 6.04 | 5.85 | 5.65 | 0.49 |
| 5 | 1.51 | 3.16 | 3.06 | 2.96 | 0.49 |
| 6 | 0.89 | 1.87 | 1.81 | 1.75 | 0.49 |

**S1 - Sample calculation**

$$d_{50,\chi,i} = d_{50,28.3,i} \times \sqrt{\frac{28.3}{\chi}}$$

$$= 7 \times \sqrt{\frac{28.3}{15}}$$

$$= 9.61\ \mu m$$

$$d_o = d_{eq} \Big/ \sqrt[3]{\frac{(C/\rho)}{(1-(\frac{RH}{100})}}$$

$$= 9.61 \Big/ \sqrt[3]{\frac{(60/1000)}{\left[1-\left(\frac{50}{100}\right)\right]}}$$

$$= 19.49\ \mu m$$

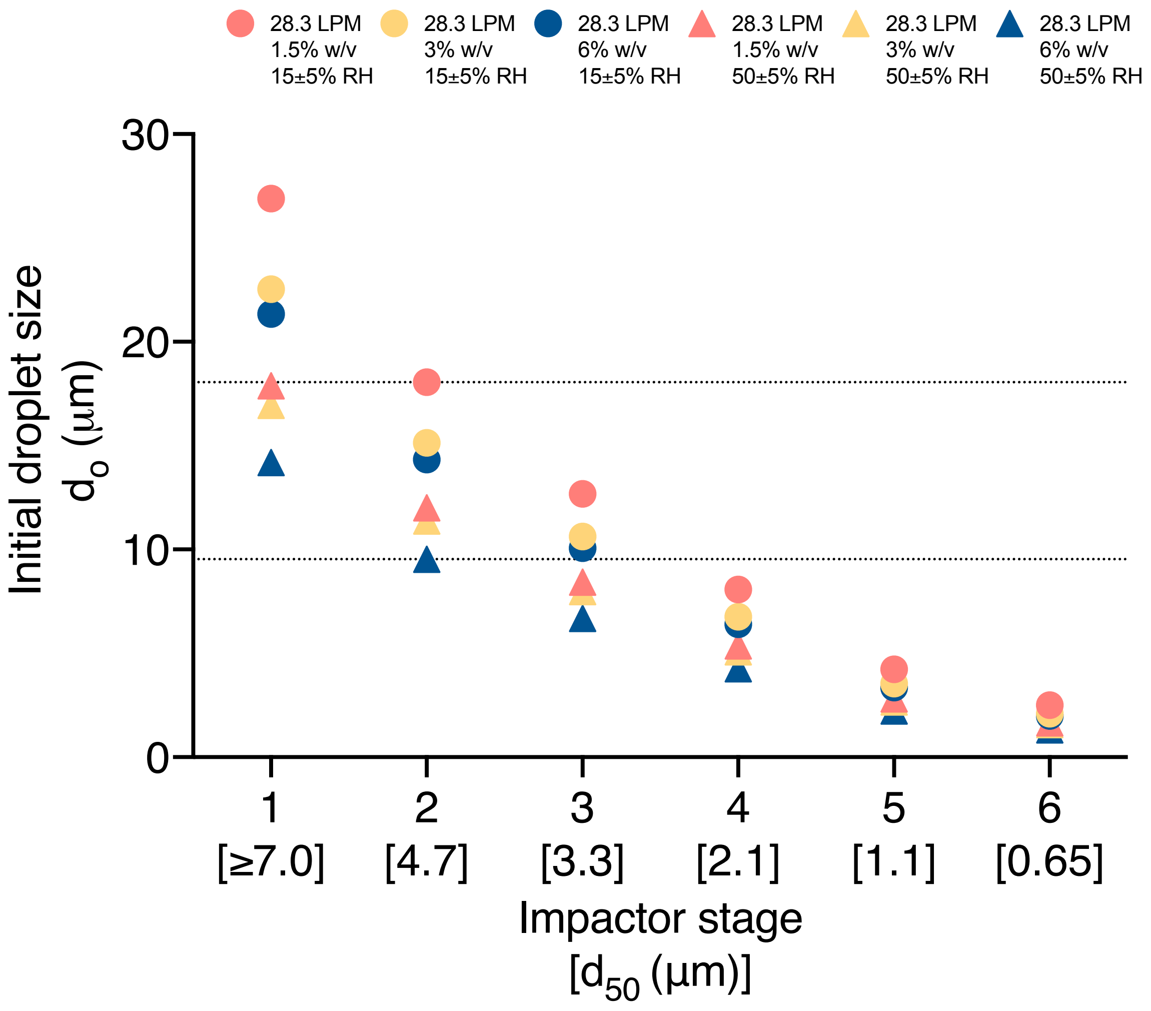


**Figure S1. Initial droplet size calculations under a standard impactor airflow rate of 28.3 LPM.** The initial size of droplets, collected below each impactor stage under the standard impactor airflow rate of 28.3 LPM, varies depending on the solute content (1.5, 3, and 6% w/v) and equilibration conditions (24±1.6°C; 15±5% or 50±5% relative humidity) within the 60-cm-high aerosol mixing chamber. The dotted lines indicate the initial size of 18.05 µm for droplets generated from 1.5% w/v peptone water solution and equilibrated at 15±5% RH and initial size of 9.53 µm for droplets generated from 6% w/v peptone water solution and equilibrated at 50±5% RH (bottom) collected below the second stage of the impactor with a 4.7 µm cut-off value under an impactor airflow rate of 28.3 LPM.

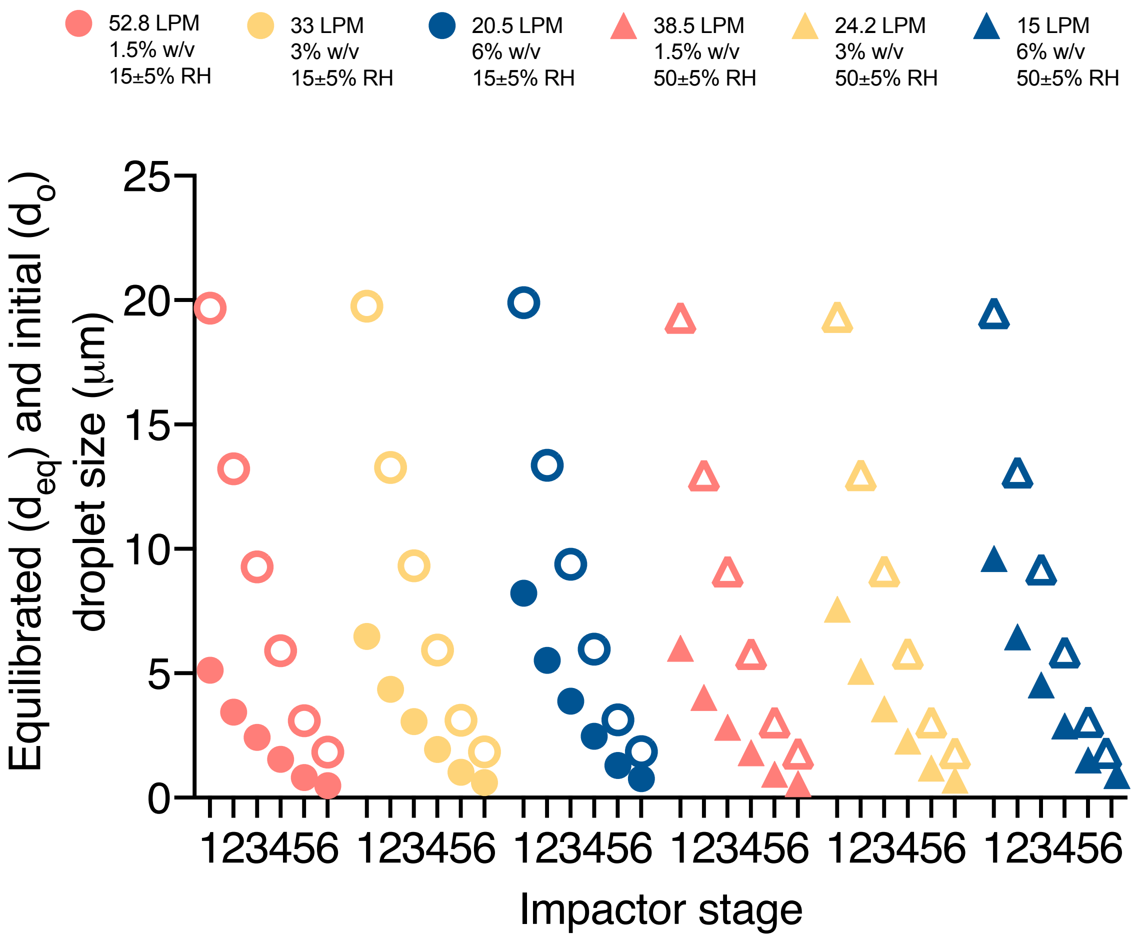


**Figure S2. Initial droplet size calculations under varied impactor airflow rates.** The graph illustrates the equilibrated (**filled shapes**) and initial (**unfilled shapes**) droplet diameters generated from 1.5% (**coral**), 3% (**yellow**), and 6% (**blue**) w/v peptone water solutions, atomized into 15±5% (**circles**) and 50±5% (**triangles**) relative humidity conditions at 24±1.6°C collected under varied impactor airflow rates ranging from 15 to 52.8 LPM.

### SECTION S2. SPECTROPHOTOMETRIC DETECTION OF 1.5, 3, AND 6% W/V PEPTONE WATER SOLUTIONS CONTAINING 1 MG/ML FLUORESCEIN, AEROSOLIZED AND COLLECTED BELOW EACH IMPACTOR STAGE UNDER 15±5% RH CONDITIONS

**Table S7. Inert aerosol droplets generated from 1.5% w/v peptone water solutions, collected under 52.8 LPM impactor airflow rate**

| Stage | $\lambda_{ex}/\lambda_{em}$, 490±5/520±5 nm | | | Avg | Percent |
|---|---|---|---|---|---|
| 1 | 3549 | 3461 | 3498 | 3503 | 2.50 |
| 2 | 3357 | 3215 | 3160 | 3244 | 2.32 |
| 3 | 3174 | 3214 | 3180 | 3189 | 2.28 |
| 4 | 1296 | 1303 | 1366 | 1322 | 0.94 |
| 5 | 8330 | 8739 | 8741 | 8603 | 6.14 |
| 6 | 9739 | 9534 | 9615 | 9629 | 6.88 |
| 1 | 2018 | 2081 | 2075 | 2058 | 1.47 |
| 2 | 2218 | 2213 | 2166 | 2199 | 1.57 |
| 3 | 1826 | 1817 | 1819 | 1821 | 1.30 |
| 4 | 1816 | 1814 | 1829 | 1820 | 1.30 |
| 5 | 919 | 848 | 810 | 859 | 0.61 |
| 6 | 1529 | 1414 | 1423 | 1455 | 1.04 |
| 1 | 2213 | 2092 | 2122 | 2142 | 1.53 |
| 2 | 3323 | 3489 | 3476 | 3429 | 2.45 |
| 3 | 3079 | 3141 | 3143 | 3121 | 2.23 |
| 4 | 1681 | 1586 | 1715 | 1661 | 1.19 |
| 5 | 1742 | 1680 | 1749 | 1724 | 1.23 |
| 6 | 2167 | 2307 | 2231 | 2235 | 1.60 |

Maximum fluorescence signal 1.5% peptone water solution containing 1 mg/mL fluorescein – 700098.75

**Table S8. Inert aerosol droplets generated from 3% w/v peptone water solutions, collected under 33 LPM impactor airflow rate**

| Stage | $\lambda_{ex}/\lambda_{em}$, 490±5/520±5 nm | | | Avg | Percent |
|---|---|---|---|---|---|
| 1 | 4501 | 4552 | 4353 | 4469 | 2.94 |
| 2 | 3787 | 3795 | 3762 | 3781 | 2.49 |
| 3 | 4144 | 3929 | 3946 | 4006 | 2.63 |
| 4 | 5808 | 5868 | 5920 | 5865 | 3.86 |
| 5 | 5414 | 5321 | 5029 | 5255 | 3.46 |
| 6 | 2262 | 2208 | 2309 | 2260 | 1.49 |
| 1 | 2349 | 2329 | 2285 | 2321 | 1.53 |
| 2 | 2358 | 2258 | 2300 | 2305 | 1.52 |
| 3 | 2251 | 2252 | 2267 | 2257 | 1.48 |
| 4 | 2864 | 2842 | 2791 | 2832 | 1.86 |
| 5 | 3300 | 3280 | 3316 | 3299 | 2.17 |
| 6 | 1713 | 1720 | 1742 | 1725 | 1.13 |
| 1 | 2549 | 2513 | 2501 | 2521 | 1.66 |
| 2 | 2748 | 2768 | 2871 | 2796 | 1.84 |
| 3 | 4142 | 4183 | 4079 | 4135 | 2.72 |
| 4 | 3128 | 3117 | 3291 | 3179 | 2.09 |
| 5 | 642 | 613 | 643 | 633 | 0.42 |
| 6 | 1347 | 1280 | 1385 | 1337 | 0.88 |

Maximum fluorescence signal 3% peptone water solution containing 1 mg/mL fluorescein – 760437.5

**Table S9. Inert aerosol droplets generated from 6% w/v peptone water solutions, collected under 20.5 LPM impactor airflow rate**

| Stage | $\lambda_{ex}/\lambda_{em}$, 490±5/520±5 nm | | | Avg | Percent |
|---|---|---|---|---|---|
| 1 | 2631 | 2586 | 2720 | 2646 | 1.69 |
| 2 | 2080 | 2067 | 2026 | 2058 | 1.32 |
| 3 | 4349 | 4462 | 4407 | 4406 | 2.82 |
| 4 | 1374 | 1412 | 1436 | 1407 | 0.90 |
| 5 | 2613 | 2863 | 2703 | 2726 | 1.75 |
| 6 | 924 | 961 | 999 | 961 | 0.62 |
| 1 | 2227 | 2276 | 2174 | 2226 | 1.43 |
| 2 | 1902 | 1904 | 1951 | 1919 | 1.23 |
| 3 | 2681 | 2679 | 2651 | 2670 | 1.71 |
| 4 | 4080 | 3969 | 3937 | 3995 | 2.56 |
| 5 | 3153 | 2985 | 3153 | 3097 | 1.98 |
| 6 | 1070 | 1016 | 1053 | 1046 | 0.67 |
| 1 | 2687 | 2639 | 2587 | 2638 | 1.69 |
| 2 | 2657 | 2719 | 2739 | 2705 | 1.73 |
| 3 | 2950 | 3050 | 3007 | 3002 | 1.92 |
| 4 | 2713 | 2792 | 2810 | 2772 | 1.78 |
| 5 | 4257 | 4251 | 4332 | 4280 | 2.74 |
| 6 | 1148 | 1209 | 1192 | 1183 | 0.76 |

Maximum fluorescence signal 3% peptone water solution containing 1 mg/mL fluorescein – 780500

### S8 - Sample calculation

$$Fluorescein\ deposition\ \ (\lambda_{ex}/\lambda_{em}, 490 \pm 5/520 \pm 5\ \text{nm}) = \left[\frac{2646 \times 5}{780500}\right] \times 100\% = 1.69\%$$

### SECTION S3. SPECTROPHOTOMETRIC DETECTION OF 1.5, 3, AND 6% W/V PEPTONE WATER SOLUTIONS CONTAINING 1 MG/ML FLUORESCEIN, AEROSOLIZED AND COLLECTED BELOW EACH IMPACTOR STAGE UNDER 50±5% RH CONDITIONS

**Table S10. Inert aerosol droplets generated from 1.5% w/v peptone water solutions, collected under 38.5 LPM impactor airflow rate**

| Stage | $\lambda_{ex}/\lambda_{em}$, 490±5/520±5 nm | | | Avg | Percent |
|---|---|---|---|---|---|
| 1 | 884 | 943 | 858 | 895 | 0.80 |
| 2 | 1216 | 1223 | 1229 | 1223 | 1.09 |
| 3 | 2093 | 2096 | 2062 | 2084 | 1.86 |
| 4 | 3361 | 3283 | 3310 | 3318 | 2.96 |
| 5 | 3883 | 3849 | 3738 | 3823 | 3.41 |
| 6 | 1070 | 1071 | 1077 | 1073 | 0.96 |
| 1 | 761 | 746 | 705 | 737 | 0.67 |
| 2 | 1169 | 1218 | 1187 | 1191 | 1.07 |
| 3 | 3783 | 3931 | 3767 | 3827 | 3.45 |
| 4 | 4169 | 4024 | 4412 | 4202 | 3.79 |
| 5 | 4576 | 4623 | 4454 | 4551 | 4.11 |
| 6 | 2189 | 2181 | 2157 | 2176 | 1.96 |
| 1 | 725 | 753 | 701 | 726 | 0.64 |
| 2 | 1316 | 1328 | 1379 | 1341 | 1.18 |
| 3 | 2524 | 2581 | 2459 | 2521 | 2.23 |
| 4 | 3197 | 3057 | 3277 | 3177 | 2.81 |
| 5 | 6011 | 5892 | 6040 | 5981 | 5.28 |
| 6 | 996 | 969 | 927 | 964 | 0.85 |

**Table S11. Inert aerosol droplets generated from 3% w/v peptone water solutions, collected under 24.2 LPM impactor airflow rate**

| Stage | $\lambda_{ex}/\lambda_{em}$, 490±5/520±5 nm | | | Avg | Percent |
|---|---|---|---|---|---|
| 1 | 3217 | 3158 | 3022 | 3132 | 2.55 |
| 2 | 2757 | 2781 | 2763 | 2767 | 2.26 |
| 3 | 3776 | 3759 | 3719 | 3751 | 3.06 |
| 4 | 5762 | 5742 | 5930 | 5811 | 4.74 |
| 5 | 8972 | 8965 | 8787 | 8908 | 7.27 |
| 6 | 1547 | 1454 | 1480 | 1494 | 1.22 |
| 1 | 1052 | 1013 | 1012 | 1026 | 0.83 |
| 2 | 645 | 680 | 664 | 663 | 0.54 |
| 3 | 1782 | 1837 | 1789 | 1803 | 1.46 |
| 4 | 1729 | 1749 | 1852 | 1777 | 1.44 |
| 5 | 2060 | 2102 | 2051 | 2071 | 1.68 |
| 6 | 896 | 887 | 875 | 886 | 0.72 |
| 1 | 691 | 587 | 594 | 624 | 0.51 |
| 2 | 845 | 771 | 809 | 808 | 0.66 |
| 3 | 566 | 566 | 544 | 559 | 0.46 |
| 4 | 1415 | 1437 | 1433 | 1428 | 1.17 |
| 5 | 2156 | 2268 | 1957 | 2127 | 1.74 |
| 6 | 736 | 725 | 700 | 720 | 0.59 |

**Table S12. Inert aerosol droplets generated from 6% w/v peptone water solutions, collected under 15 LPM impactor airflow rate**

| Stage | $\lambda_{ex}/\lambda_{em}$, 490±5/520±5 nm | | | Avg | Percent |
|---|---|---|---|---|---|
| 1 | 2512 | 2530 | 2431 | 2491 | 1.76 |
| 2 | 1472 | 1434 | 1467 | 1458 | 1.03 |
| 3 | 1985 | 2026 | 2047 | 2019 | 1.43 |
| 4 | 2862 | 2890 | 2850 | 2867 | 2.03 |
| 5 | 5901 | 5844 | 5915 | 5887 | 4.17 |
| 6 | 1746 | 1796 | 1730 | 1757 | 1.24 |
| 1 | 901 | 816 | 834 | 850 | 0.60 |
| 2 | 693 | 744 | 747 | 728 | 0.51 |
| 3 | 1163 | 1129 | 1107 | 1133 | 0.80 |
| 4 | 1691 | 1864 | 1711 | 1755 | 1.24 |
| 5 | 2299 | 2271 | 2253 | 2274 | 1.61 |
| 6 | 1007 | 1081 | 1059 | 1049 | 0.74 |
| 1 | 1664 | 1638 | 1581 | 1628 | 1.18 |
| 2 | 938 | 1026 | 985 | 983 | 0.71 |
| 3 | 1653 | 1703 | 1693 | 1683 | 1.22 |
| 4 | 3955 | 3933 | 4075 | 3988 | 2.90 |
| 5 | 5018 | 5016 | 4953 | 4996 | 3.63 |
| 6 | 1879 | 1933 | 1920 | 1911 | 1.39 |

### SECTION S4. ONE-WAY ANOVA FOLLOWED BY TUKEY'S MULTIPLE COMPARISONS TEST RESULTS FOR DOSES OF 1.5, 3, AND 6% W/V PEPTONE WATER SOLUTIONS CONTAINING 1 MG/ML FLUORESCEIN AEROSOLIZED AND COLLECTED BELOW EACH IMPACTOR STAGE

**Table S13. Percent nominal dose of sodium fluorescein collected below the first impactor stage**

| Tukey's multiple comparisons test | Summary | Adjusted P Value |
|---|---|---|
| 1.5% w/v 15±5% RH vs. 3% w/v 15±5% RH | ns | 0.9985 |
| 1.5% w/v 15±5% RH vs. 6% RH 15±5% RH | ns | 0.9975 |
| 1.5% w/v 15±5% RH vs. 1.5% w/v 50±5% RH | ns | 0.3307 |
| 1.5% w/v 15±5% RH vs. 3% w/v 50±5% RH | ns | 0.9057 |
| 1.5% w/v 15±5% RH vs. 6% RH 50±5% RH | ns | 0.8148 |
| 3% w/v 15±5% RH vs. 6% RH 15±5% RH | ns | 0.9568 |
| 3% w/v 15±5% RH vs. 1.5% w/v 50±5% RH | ns | 0.1898 |
| 3% w/v 15±5% RH vs. 3% w/v 50±5% RH | ns | 0.7260 |
| 3% w/v 15±5% RH vs. 6% RH 50±5% RH | ns | 0.6016 |
| 6% RH 15±5% RH vs. 1.5% w/v 50±5% RH | ns | 0.5539 |
| 6% RH 15±5% RH vs. 3% w/v 50±5% RH | ns | 0.9909 |
| 6% RH 15±5% RH vs. 6% RH 50±5% RH | ns | 0.9632 |
| 1.5% w/v 50±5% RH vs. 3% w/v 50±5% RH | ns | 0.8605 |
| 1.5% w/v 50±5% RH vs. 6% RH 50±5% RH | ns | 0.9367 |
| 3% w/v 50±5% RH vs. 6% RH 50±5% RH | ns | >0.9999 |

**Table S14. Percent nominal dose of sodium fluorescein collected below the second impactor stage**

| Tukey's multiple comparisons test | Summary | Adjusted P Value |
|---|---|---|
| 1.5% w/v 15±5% RH vs. 3% w/v 15±5% RH | ns | 0.9983 |
| 1.5% w/v 15±5% RH vs. 6% RH 15±5% RH | ns | 0.5783 |
| 1.5% w/v 15±5% RH vs. 1.5% w/v 50±5% RH | ns | 0.2256 |
| 1.5% w/v 15±5% RH vs. 3% w/v 50±5% RH | ns | 0.2555 |
| 1.5% w/v 15±5% RH vs. 6% RH 50±5% RH | ns | 0.0559 |
| 3% w/v 15±5% RH vs. 6% RH 15±5% RH | ns | 0.7999 |
| 3% w/v 15±5% RH vs. 1.5% w/v 50±5% RH | ns | 0.3894 |
| 3% w/v 15±5% RH vs. 3% w/v 50±5% RH | ns | 0.4327 |
| 3% w/v 15±5% RH vs. 6% RH 50±5% RH | ns | 0.1080 |
| 6% RH 15±5% RH vs. 1.5% w/v 50±5% RH | ns | 0.9707 |
| 6% RH 15±5% RH vs. 3% w/v 50±5% RH | ns | 0.9826 |
| 6% RH 15±5% RH vs. 6% RH 50±5% RH | ns | 0.5957 |
| 1.5% w/v 50±5% RH vs. 3% w/v 50±5% RH | ns | >0.9999 |
| 1.5% w/v 50±5% RH vs. 6% RH 50±5% RH | ns | 0.9447 |
| 3% w/v 50±5% RH vs. 6% RH 50±5% RH | ns | 0.9205 |

**Table S15. Percent nominal dose of sodium fluorescein collected below the third impactor stage**

| Tukey's multiple comparisons test | Summary | Adjusted P Value |
|---|---|---|
| 1.5% w/v 15±5% RH vs. 3% w/v 15±5% RH | ns | 0.9932 |
| 1.5% w/v 15±5% RH vs. 6% RH 15±5% RH | ns | 0.9992 |
| 1.5% w/v 15±5% RH vs. 1.5% w/v 50±5% RH | ns | 0.9376 |
| 1.5% w/v 15±5% RH vs. 3% w/v 50±5% RH | ns | 0.9975 |
| 1.5% w/v 15±5% RH vs. 6% RH 50±5% RH | ns | 0.8141 |
| 3% w/v 15±5% RH vs. 6% RH 15±5% RH | ns | >0.9999 |
| 3% w/v 15±5% RH vs. 1.5% w/v 50±5% RH | ns | 0.9989 |
| 3% w/v 15±5% RH vs. 3% w/v 50±5% RH | ns | 0.9180 |
| 3% w/v 15±5% RH vs. 6% RH 50±5% RH | ns | 0.5166 |
| 6% RH 15±5% RH vs. 1.5% w/v 50±5% RH | ns | 0.9916 |
| 6% RH 15±5% RH vs. 3% w/v 50±5% RH | ns | 0.9669 |
| 6% RH 15±5% RH vs. 6% RH 50±5% RH | ns | 0.6300 |
| 1.5% w/v 50±5% RH vs. 3% w/v 50±5% RH | ns | 0.7590 |
| 1.5% w/v 50±5% RH vs. 6% RH 50±5% RH | ns | 0.3316 |
| 3% w/v 50±5% RH vs. 6% RH 50±5% RH | ns | 0.9627 |

**Table S16. Percent nominal dose of sodium fluorescein collected below the fourth impactor stage**

| Tukey's multiple comparisons test | Summary | Adjusted P Value |
|---|---|---|
| 1.5% w/v 15±5% RH vs. 3% w/v 15±5% RH | ns | 0.5701 |
| 1.5% w/v 15±5% RH vs. 6% RH 15±5% RH | ns | 0.9795 |
| 1.5% w/v 15±5% RH vs. 1.5% w/v 50±5% RH | ns | 0.2494 |
| 1.5% w/v 15±5% RH vs. 3% w/v 50±5% RH | ns | 0.6709 |
| 1.5% w/v 15±5% RH vs. 6% RH 50±5% RH | ns | 0.8920 |
| 3% w/v 15±5% RH vs. 6% RH 15±5% RH | ns | 0.9145 |
| 3% w/v 15±5% RH vs. 1.5% w/v 50±5% RH | ns | 0.9823 |
| 3% w/v 15±5% RH vs. 3% w/v 50±5% RH | ns | >0.9999 |
| 3% w/v 15±5% RH vs. 6% RH 50±5% RH | ns | 0.9867 |
| 6% RH 15±5% RH vs. 1.5% w/v 50±5% RH | ns | 0.5834 |
| 6% RH 15±5% RH vs. 3% w/v 50±5% RH | ns | 0.9604 |
| 6% RH 15±5% RH vs. 6% RH 50±5% RH | ns | 0.9990 |
| 1.5% w/v 50±5% RH vs. 3% w/v 50±5% RH | ns | 0.9529 |
| 1.5% w/v 50±5% RH vs. 6% RH 50±5% RH | ns | 0.7826 |
| 3% w/v 50±5% RH vs. 6% RH 50±5% RH | ns | 0.9970 |

**Table S17. Percent nominal dose of sodium fluorescein collected below the fifth impactor stage**

| Tukey's multiple comparisons test | Summary | Adjusted P Value |
|---|---|---|
| 1.5% w/v 15±5% RH vs. 3% w/v 15±5% RH | ns | 0.9985 |
| 1.5% w/v 15±5% RH vs. 6% RH 15±5% RH | ns | 0.9995 |
| 1.5% w/v 15±5% RH vs. 1.5% w/v 50±5% RH | ns | 0.9203 |
| 1.5% w/v 15±5% RH vs. 3% w/v 50±5% RH | ns | 0.9931 |
| 1.5% w/v 15±5% RH vs. 6% RH 50±5% RH | ns | 0.9997 |
| 3% w/v 15±5% RH vs. 6% RH 15±5% RH | ns | >0.9999 |
| 3% w/v 15±5% RH vs. 1.5% w/v 50±5% RH | ns | 0.7499 |
| 3% w/v 15±5% RH vs. 3% w/v 50±5% RH | ns | 0.9301 |
| 3% w/v 15±5% RH vs. 6% RH 50±5% RH | ns | 0.9813 |
| 6% RH 15±5% RH vs. 1.5% w/v 50±5% RH | ns | 0.7949 |
| 6% RH 15±5% RH vs. 3% w/v 50±5% RH | ns | 0.9524 |
| 6% RH 15±5% RH vs. 6% RH 50±5% RH | ns | 0.9898 |
| 1.5% w/v 50±5% RH vs. 3% w/v 50±5% RH | ns | 0.9978 |
| 1.5% w/v 50±5% RH vs. 6% RH 50±5% RH | ns | 0.9809 |
| 3% w/v 50±5% RH vs. 6% RH 50±5% RH | ns | 0.9998 |

**Table S18. Percent nominal dose of sodium fluorescein collected below the first impactor stage**

| Tukey's multiple comparisons test | Summary | Adjusted P Value |
|---|---|---|
| 1.5% w/v 15±5% RH vs. 3% w/v 15±5% RH | ns | 0.4965 |
| 1.5% w/v 15±5% RH vs. 6% RH 15±5% RH | ns | 0.2870 |
| 1.5% w/v 15±5% RH vs. 1.5% w/v 50±5% RH | ns | 0.5420 |
| 1.5% w/v 15±5% RH vs. 3% w/v 50±5% RH | ns | 0.3487 |
| 1.5% w/v 15±5% RH vs. 6% RH 50±5% RH | ns | 0.4762 |
| 3% w/v 15±5% RH vs. 6% RH 15±5% RH | ns | 0.9974 |
| 3% w/v 15±5% RH vs. 1.5% w/v 50±5% RH | ns | >0.9999 |
| 3% w/v 15±5% RH vs. 3% w/v 50±5% RH | ns | 0.9996 |
| 3% w/v 15±5% RH vs. 6% RH 50±5% RH | ns | >0.9999 |
| 6% RH 15±5% RH vs. 1.5% w/v 50±5% RH | ns | 0.9943 |
| 6% RH 15±5% RH vs. 3% w/v 50±5% RH | ns | >0.9999 |
| 6% RH 15±5% RH vs. 6% RH 50±5% RH | ns | 0.9983 |
| 1.5% w/v 50±5% RH vs. 3% w/v 50±5% RH | ns | 0.9988 |
| 1.5% w/v 50±5% RH vs. 6% RH 50±5% RH | ns | >0.9999 |
| 3% w/v 50±5% RH vs. 6% RH 50±5% RH | ns | 0.9998 |

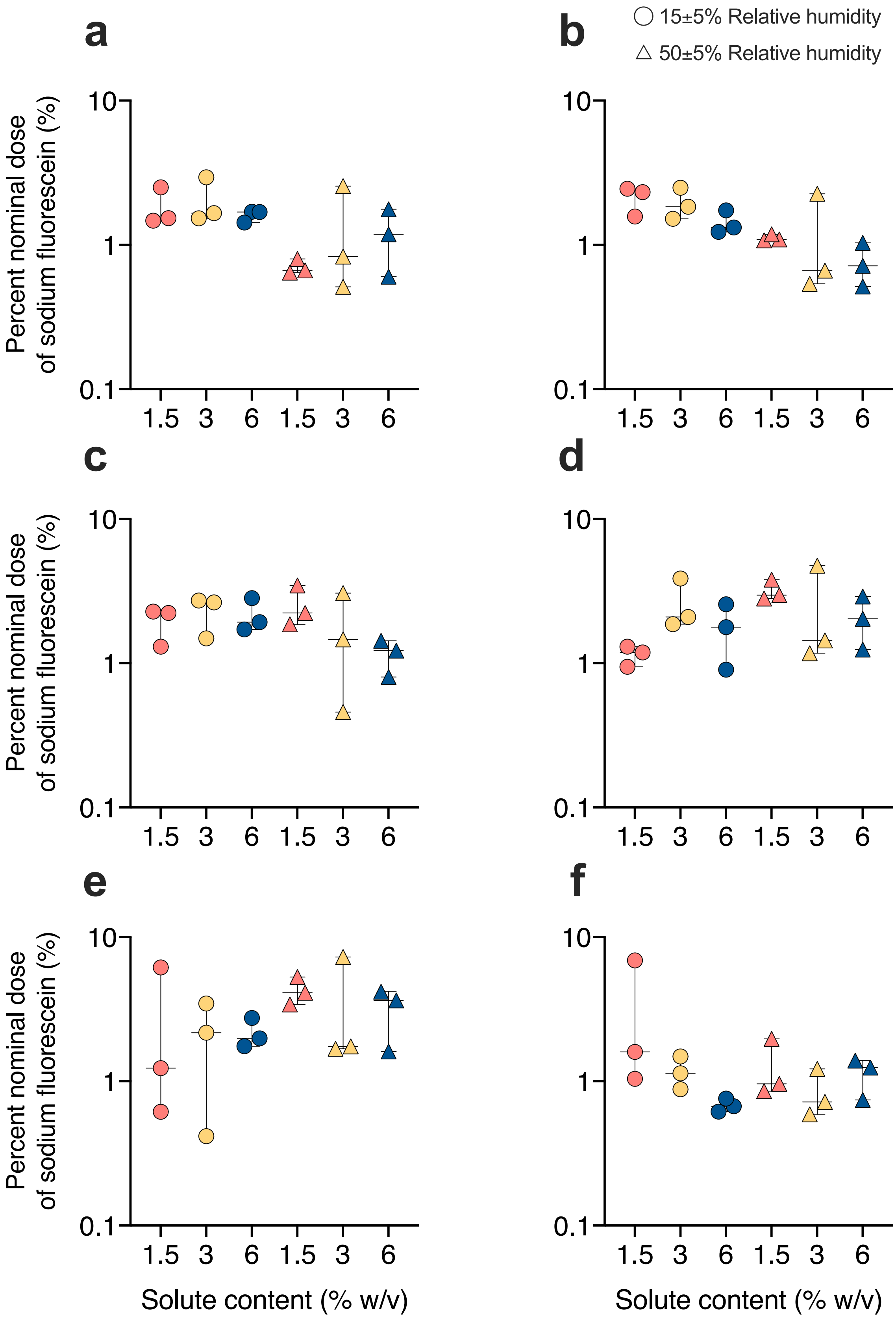
a
b
c
d
e
f
15±5% Relative humidity
50±5% Relative humidity
Percent nominal dose of sodium fluorescein (%)
10
1
0.1
1.5
3
6
Solute content (% w/v)

**Figure S3. Effect of solute content, relative humidity, and impactor airflow rates on the nominal dose of fluorescein-laden droplets collected below each impactor stage.** Solutions containing 1 mg/mL sodium fluorescein, prepared in 1.5% (**coral**), 3% (**yellow**), and 6% (**blue**) w/v peptone water solutions, were atomized into 15±5% (**circles**) and 50±5% (**triangles**) relative humidity conditions at 24±1.6°C. The graphs present the nominal dose percentage recovered from stages (**a**) 1, (**b**) 2, (**c**) 3, (**d**) 4, (**e**) 5, and (**f**) 6. Data from three replicate trials are presented (n=3). Spectrophotometric measurements of fluorescein wash samples were performed under excitation and emission wavelengths of 490±5 and 515±5 nm, respectively. Doses recovered below each impactor stage across the six conditions, analysed using two-way ANOVA followed by Tukey's multiple comparisons, were found to be non-significant, ns - $p>0.05$.

### SECTION S5. RECOVERED COUNTS OF VIABLE PATHOGEN-LADEN DROPLETS GENERATED FROM 1×10$^5$ PFU/ML PHAGE SUSPENSION COLLECTED UNDER 15±5% RH

$N_R$ = Raw counts; $N_C$ = Coincidence-correct counts; MDS = Mean droplet size (µm)

**Table S19. Viable pathogen-laden droplet counts generated from 1.5% w/v peptone water suspension, collected under 52.8 LPM impactor airflow rate**

| $d_{50}$ | $N_R$ | $N_C$ | $N_R$ | $N_C$ | $N_R$ | $N_C$ | $N_R$ | $N_C$ |
|---|---|---|---|---|---|---|---|---|
| 5.12 | 118 | 140 | 77 | 86 | 174 | 228 | 177 | 234 |
| 3.44 | 262 | 426 | 167 | 216 | 207 | 292 | 305 | 575 |
| 2.42 | 294 | 531 | 250 | 392 | 300 | 555 | 303 | 567 |
| 1.54 | 311 | 601 | 335 | 727 | 263 | 429 | 307 | 584 |
| 0.81 | 179 | 237 | 302 | 563 | 207 | 292 | 212 | 302 |
| 0.48 | 71 | 78 | 139 | 171 | 69 | 76 | 73 | 81 |
| Total | | 2013 | | 2155 | | 1872 | | 2343 |
| MDS | | 2.29 | | 1.76 | | 2.37 | | 2.44 |

**Table S20. Dataset 1- Viable pathogen-laden droplet counts generated from 3% w/v peptone water suspension, collected under 33 LPM impactor airflow rate**

| $d_{50}$ | $N_R$ | $N_C$ | $N_R$ | $N_C$ | $N_R$ | $N_C$ |
|---|---|---|---|---|---|---|
| 6.48 | 136 | 166 | 165 | 213 | 80 | 89 |
| 4.35 | 188 | 254 | 216 | 311 | 105 | 122 |
| 3.06 | 255 | 406 | 241 | 369 | 189 | 256 |
| 1.94 | 249 | 390 | 307 | 584 | 237 | 359 |
| 1.02 | 248 | 387 | 193 | 264 | 238 | 362 |
| 0.60 | 41 | 43 | 36 | 38 | 98 | 112 |
| Total | | 1646 | | 1779 | | 1300 |
| MPS | | 2.80 | | 2.97 | | 2.33 |

**Table S21. Dataset 2 - Viable aerosol droplets generated from 3% w/v peptone water suspension, collected under 33 LPM impactor airflow rate**

| $d_{50}$ | $N_R$ | $N_C$ | $N_R$ | $N_C$ | $N_R$ | $N_C$ |
|---|---|---|---|---|---|---|
| 6.48 | 107 | 125 | 167 | 216 | 196 | 269 |
| 4.35 | 161 | 206 | 232 | 347 | 253 | 400 |
| 3.06 | 217 | 313 | 297 | 543 | 268 | 444 |
| 1.94 | 219 | 317 | 299 | 551 | 285 | 499 |
| 1.02 | 169 | 220 | 235 | 354 | 205 | 287 |
| 0.60 | 48 | 51 | 68 | 75 | 57 | 62 |
| Total | | 1232 | | 2086 | | 1961 |
| MPS | | 2.87 | | 2.90 | | 3.13 |

**Table S22. Dataset 1 - Viable pathogen-laden droplet counts generated from 6% w/v peptone water suspension, collected under 20.5 LPM impactor airflow rate**

| $d_{50}$ | $N_R$ | $N_C$ | $N_R$ | $N_C$ | $N_R$ | $N_C$ |
|---|---|---|---|---|---|---|
| 8.22 | 154 | 194 | 60 | 65 | 77 | 86 |
| 5.52 | 155 | 196 | 51 | 55 | 99 | 114 |
| 3.88 | 230 | 342 | 88 | 99 | 157 | 199 |
| 2.47 | 239 | 364 | 117 | 138 | 186 | 250 |
| 1.29 | 228 | 338 | 116 | 137 | 152 | 191 |
| 0.76 | 73 | 81 | 29 | 30 | 22 | 23 |
| Total | | 1515 | | 524 | | 863 |
| MPS | | 3.56 | | 3.36 | | 3.46 |

**Table S23. Dataset 2 - Viable pathogen-laden droplet counts generated from 6% w/v peptone water suspension, collected under 20.5 LPM impactor airflow rate**

| $d_{50}$ | $N_R$ | $N_C$ | $N_R$ | $N_C$ |
|---|---|---|---|---|
| 8.22 | 89 | 101 | 165 | 213 |
| 5.52 | 86 | 97 | 143 | 177 |
| 3.88 | 158 | 201 | 217 | 313 |
| 2.47 | 210 | 298 | 217 | 313 |
| 1.29 | 186 | 250 | 216 | 311 |
| 0.76 | 55 | 59 | 69 | 76 |
| Total | | 1006 | | 1403 |
| MPS | | 3.23 | | 3.69 |

### SECTION S6. RECOVERED COUNTS OF VIABLE PATHOGEN-LADEN DROPLETS GENERATED FROM 1×$10^5$ PFU/ML PHAGE SUSPENSION COLLECTED UNDER 50±5% RH

**Table S24. Dataset 1 - Viable pathogen-laden droplet counts generated from 1.5% w/v peptone water suspension, collected under 38.5 LPM impactor airflow rate**

| $d_{50}$ | $N_R$ | $N_C$ | $N_R$ | $N_C$ | $N_R$ | $N_C$ | $N_R$ | $N_C$ |
|---|---|---|---|---|---|---|---|---|
| 6.00 | 63 | 69 | 123 | 147 | 113 | 133 | 51 | 55 |
| 4.03 | 118 | 140 | 182 | 243 | 217 | 313 | 90 | 102 |
| 2.83 | 140 | 172 | 256 | 409 | 308 | 588 | 148 | 185 |
| 1.80 | 134 | 163 | 283 | 492 | 301 | 559 | 176 | 232 |
| 0.94 | 96 | 110 | 192 | 262 | 233 | 349 | 126 | 151 |
| 0.56 | 20 | 21 | 56 | 60 | 48 | 51 | 17 | 17 |
| Total | | 675 | | 1613 | | 1993 | | 742 |
| MPS | | 2.78 | | 2.59 | | 2.55 | | 2.47 |

**Table S25. Dataset 2 – Viable pathogen-laden droplet counts generated from 1.5% w/v peptone water suspension, collected under 38.5 LPM impactor airflow rate**

| $d_{50}$ | $N_R$ | $N_C$ | $N_R$ | $N_C$ | $N_R$ | $N_C$ |
|---|---|---|---|---|---|---|
| 6.00 | 96 | 110 | 89 | 101 | 93 | 106 |
| 4.03 | 182 | 243 | 185 | 248 | 198 | 273 |
| 2.83 | 239 | 364 | 232 | 347 | 244 | 377 |
| 1.80 | 239 | 364 | 206 | 289 | 250 | 392 |
| 0.94 | 188 | 254 | 159 | 203 | 186 | 250 |
| 0.56 | 24 | 25 | 25 | 26 | 21 | 22 |
| Total | | 1360 | | 1214 | | 1420 |
| MPS | | 2.63 | | 2.73 | | 2.65 |

**Table S26. Dataset 1 - Viable pathogen-laden droplet counts generated from 3% w/v peptone water suspension, collected under 24.2 LPM impactor airflow rate**

| $d_{50}$ | $N_R$ | $N_C$ | $N_R$ | $N_C$ | $N_R$ | $N_C$ | $N_R$ | $N_C$ |
|---|---|---|---|---|---|---|---|---|
| 7.57 | 62 | 67 | 145 | 180 | 166 | 214 | 97 | 111 |
| 5.08 | 176 | 232 | 204 | 285 | 168 | 218 | 82 | 92 |
| 3.57 | 173 | 227 | 256 | 409 | 285 | 499 | 174 | 228 |
| 2.27 | 165 | 213 | 259 | 417 | 252 | 398 | 197 | 271 |
| 1.19 | 98 | 112 | 196 | 269 | 250 | 392 | 150 | 188 |
| 0.70 | 6 | 6 | 53 | 57 | 42 | 44 | 30 | 31 |
| Total | | 857 | | 1617 | | 1765 | | 921 |
| MPS | | 3.64 | | 3.45 | | 3.35 | | 3.24 |

**Table S27. Dataset 2 - Viable pathogen-laden droplet counts generated from 1×10$^5$ PFU/mL phage suspension in 3% w/v peptone water, collected under 24.2 LPM impactor airflow rate**

| $d_{50}$ | $N_R$ | $N_C$ | $N_R$ | $N_C$ | $N_R$ | $N_C$ |
|---|---|---|---|---|---|---|
| 7.57 | 88 | 99 | 115 | 136 | 136 | 166 |
| 5.08 | 125 | 150 | 186 | 250 | 188 | 254 |
| 3.57 | 169 | 220 | 199 | 275 | 219 | 317 |
| 2.27 | 195 | 267 | 203 | 283 | 262 | 426 |
| 1.19 | 165 | 213 | 164 | 211 | 201 | 279 |
| 0.70 | 36 | 38 | 43 | 46 | 56 | 60 |
| Total | | 987 | | 1201 | | 1502 |
| MPS | | 3.23 | | 3.50 | | 3.34 |

**Table S28. Dataset 1 - Viable pathogen-laden droplet counts generated from 6% w/v peptone water suspension, collected under 15 LPM impactor airflow rate**

| $d_{50}$ | $N_R$ | $N_C$ | $N_R$ | $N_C$ | $N_R$ | $N_C$ | $N_R$ | $N_C$ |
|---|---|---|---|---|---|---|---|---|
| 9.61 | 109 | 127 | 54 | 58 | 140 | 172 | 86 | 97 |
| 6.46 | 162 | 208 | 63 | 69 | 104 | 120 | 73 | 81 |
| 4.53 | 204 | 285 | 75 | 83 | 129 | 156 | 106 | 123 |
| 2.88 | 138 | 169 | 150 | 188 | 165 | 213 | 147 | 183 |
| 1.51 | 114 | 134 | 163 | 209 | 130 | 157 | 144 | 179 |
| 0.89 | 19 | 19 | 40 | 42 | 44 | 47 | 52 | 56 |
| Total | | 942 | | 649 | | 865 | | 719 |
| MPS | | 4.84 | | 3.51 | | 4.66 | | 3.98 |

**Table S29. Dataset 2 - Viable pathogen-laden droplet counts generated from 6% w/v peptone water suspension, collected under 15 LPM impactor airflow rate**

| $d_{50}$ | $N_R$ | $N_C$ | $N_R$ | $N_C$ | $N_R$ | $N_C$ |
|---|---|---|---|---|---|---|
| 9.61 | 83 | 93 | 157 | 199 | 142 | 175 |
| 6.46 | 58 | 63 | 110 | 129 | 107 | 125 |
| 4.53 | 97 | 111 | 162 | 208 | 174 | 228 |
| 2.88 | 109 | 127 | 205 | 287 | 197 | 271 |
| 1.51 | 74 | 82 | 176 | 232 | 183 | 245 |
| 0.89 | 19 | 19 | 61 | 66 | 50 | 53 |
| Total | | 495 | | 1121 | | 1097 |
| MPS | | 4.67 | | 4.39 | | 4.30 |

### SECTION S7. RECOVERED COUNTS OF VIABLE PATHOGEN-LADEN DROPLETS GENERATED FROM 5×10$^5$ PFU/ML PHAGE SUSPENSION COLLECTED UNDER 15±5% RH

**Table S30. Viable pathogen-laden droplet counts generated from 1.5% w/v peptone water suspension, collected under 52.8 LPM impactor airflow rate**

| $d_{50}$ | $N_R$ | $N_C$ | $N_R$ | $N_C$ | $N_R$ | $N_C$ | $N_R$ | $N_C$ |
|---|---|---|---|---|---|---|---|---|
| 5.12 | 254 | 403 | 243 | 374 | 262 | 426 | 238 | 362 |
| 3.44 | 353 | 857 | 341 | 766 | 384 | 1288 | 379 | 1179 |
| 2.42 | 361 | 931 | 386 | 1341 | 393 | 1619 | 398 | 2128 |
| 1.54 | 342 | 772 | 383 | 1263 | 398 | 2128 | 397 | 1961 |
| 0.81 | 237 | 359 | 296 | 539 | 396 | 1844 | 361 | 931 |
| 0.48 | 80 | 89 | 110 | 129 | 202 | 281 | 205 | 287 |
| Total | | 3411 | | 4412 | | 7586 | | 6848 |
| MPS | | 2.57 | | 2.32 | | 2.03 | | 2.18 |

**Table S31. Viable pathogen-laden droplet counts generated from 3% w/v peptone water suspension, collected under 33 LPM impactor airflow rate**

| $d_{50}$ | $N_R$ | $N_C$ | $N_R$ | $N_C$ | $N_R$ | $N_C$ | $N_R$ | $N_C$ | $N_R$ | $N_C$ |
|---|---|---|---|---|---|---|---|---|---|---|
| 6.48 | 265 | 435 | 249 | 390 | 316 | 624 | 276 | 469 | 278 | 475 |
| 4.35 | 332 | 709 | 321 | 649 | 392 | 1565 | 359 | 911 | 339 | 752 |
| 3.06 | 378 | 1160 | 382 | 1241 | 399 | 2428 | 390 | 1476 | 393 | 1619 |
| 1.94 | 389 | 1438 | 364 | 963 | 400 | 2628 | 400 | 2628 | 399 | 2428 |
| 1.02 | 324 | 664 | 325 | 670 | 398 | 2128 | 389 | 1438 | 391 | 1518 |
| 0.60 | 82 | 92 | 87 | 98 | 248 | 387 | 228 | 338 | 226 | 333 |
| Total | | 4498 | | 4011 | | 9760 | | 7260 | | 7125 |
| MPS | | 2.89 | | 2.93 | | 2.64 | | 2.52 | | 2.49 |

**Table S32. Viable pathogen-laden droplet counts generated from 6% w/v peptone water suspension, collected under 20.5 LPM impactor airflow rate**

| $d_{50}$ | $N_R$ | $N_C$ | $N_R$ | $N_C$ | $N_R$ | $N_C$ |
|---|---|---|---|---|---|---|
| 8.22 | 270 | 450 | 306 | 579 | 272 | 456 |
| 5.52 | 277 | 472 | 312 | 606 | 246 | 382 |
| 3.88 | 340 | 759 | 374 | 1093 | 382 | 1241 |
| 2.47 | 371 | 1050 | 379 | 1179 | 397 | 1961 |
| 1.29 | 346 | 801 | 348 | 816 | 386 | 1341 |
| 0.76 | 137 | 168 | 137 | 168 | 238 | 362 |
| Total | | 3700 | | 4441 | | 5743 |
| MPS | | 3.51 | | 3.70 | | 3.05 |

**Table S33. Viable pathogen-laden droplet counts generated from 6% w/v peptone water suspension, collected under 20.5 LPM impactor airflow rate**

| $d_{50}$ | $N_R$ | $N_C$ | $N_R$ | $N_C$ | $N_R$ | $N_C$ |
|---|---|---|---|---|---|---|
| 8.22 | 257 | 411 | 248 | 387 | 281 | 485 |
| 5.52 | 246 | 382 | 267 | 440 | 338 | 746 |
| 3.88 | 346 | 801 | 351 | 840 | 392 | 1565 |
| 2.47 | 396 | 1844 | 390 | 1476 | 395 | 1754 |
| 1.29 | 387 | 1371 | 384 | 1288 | 398 | 2128 |
| 0.76 | 227 | 335 | 257 | 411 | 285 | 1314 |
| Total | | 5144 | | 4842 | | 7992 |
| MPS | | 2.95 | | 2.99 | | 2.79 |

### SECTION S8. RECOVERED COUNTS OF VIABLE PATHOGEN-LADEN DROPLETS GENERATED FROM 5×10$^5$ PFU/ML PHAGE SUSPENSION COLLECTED UNDER 50±5% RH

**Table S34. Viable pathogen-laden droplet counts generated from 1.5% w/v peptone water suspension, collected under 38.5 LPM impactor airflow rate**

| $d_{50}$ | $N_R$ | $N_C$ | $N_R$ | $N_C$ | $N_R$ | $N_C$ | $N_R$ | $N_C$ | $N_R$ | $N_C$ |
|---|---|---|---|---|---|---|---|---|---|---|
| 6.00 | 256 | 409 | 245 | 379 | 283 | 492 | 137 | 168 | 262 | 426 |
| 4.03 | 364 | 963 | 382 | 1241 | 398 | 2128 | 236 | 357 | 360 | 921 |
| 2.83 | 396 | 1844 | 393 | 1619 | 400 | 2628 | 333 | 715 | 393 | 1619 |
| 1.80 | 395 | 1754 | 393 | 1619 | 400 | 2628 | 369 | 1023 | 382 | 1241 |
| 0.94 | 380 | 1198 | 379 | 1179 | 398 | 2128 | 376 | 1125 | 366 | 986 |
| 0.56 | 159 | 203 | 158 | 201 | 226 | 333 | 171 | 223 | 113 | 133 |
| Total | | 6371 | | 6238 | | 10337 | | 3611 | | 5326 |
| MPS | | 2.50 | | 2.56 | | 2.50 | | 2.08 | | 2.64 |

**Table S35. Viable pathogen-laden droplet counts generated from 3% w/v peptone water suspension, collected under 24.2 LPM impactor airflow rate**

| $d_{50}$ | $N_R$ | $N_C$ | $N_R$ | $N_C$ | $N_R$ | $N_C$ | $N_R$ | $N_C$ | $N_R$ | $N_C$ |
|---|---|---|---|---|---|---|---|---|---|---|
| 7.57 | 274 | 462 | 300 | 555 | 283 | 492 | 291 | 520 | 300 | 555 |
| 5.08 | 330 | 697 | 372 | 1064 | 348 | 816 | 347 | 809 | 307 | 584 |
| 3.57 | 369 | 1023 | 398 | 2128 | 377 | 1142 | 382 | 1241 | 370 | 1036 |
| 2.27 | 379 | 1179 | 397 | 1961 | 398 | 2128 | 389 | 1438 | 384 | 1288 |
| 1.19 | 382 | 1241 | 389 | 1438 | 387 | 1371 | 379 | 1179 | 377 | 1142 |
| 0.70 | 161 | 206 | 218 | 315 | 203 | 283 | 162 | 208 | 109 | 127 |
| Total | | 4808 | | 7461 | | 6232 | | 5395 | | 4732 |
| MDS | | 3.12 | | 3.16 | | 2.99 | | 3.21 | | 3.22 |

**Table S36. Viable pathogen-laden droplet counts generated from 6% w/v peptone water suspension, collected under 15 LPM impactor airflow rate**

| $d_{50}$ | $N_R$ | $N_C$ | $N_R$ | $N_C$ | $N_R$ | $N_C$ | $N_R$ | $N_C$ | $N_R$ | $N_C$ |
|---|---|---|---|---|---|---|---|---|---|---|
| 9.61 | 333 | 715 | 338 | 746 | 343 | 779 | 305 | 575 | 271 | 453 |
| 6.46 | 284 | 495 | 302 | 563 | 321 | 649 | 243 | 374 | 253 | 400 |
| 4.53 | 344 | 786 | 329 | 692 | 376 | 1125 | 331 | 703 | 312 | 606 |
| 2.88 | 369 | 1023 | 376 | 1125 | 386 | 1341 | 358 | 902 | 358 | 902 |
| 1.51 | 374 | 1093 | 382 | 1241 | 396 | 1844 | 342 | 772 | 335 | 727 |
| 0.89 | 174 | 228 | 202 | 281 | 224 | 328 | 145 | 180 | 117 | 138 |
| Total | | 4340 | | 4648 | | 6066 | | 3506 | | 3226 |
| MDS | | 4.25 | | 4.16 | | 3.91 | | 4.30 | | 4.19 |

### SECTION S9. NORMALITY TEST RESULTS FOR RECOVERED COUNTS OF VIABLE PATHOGEN-LADEN DROPLETS

The parametric statistical analysis requires datasets to follow a normal distribution and is typically applied to datasets with n≥10 for the outcome of the statistical analysis to be reliable. The parametric statistical analysis in this study is justified because all 76 replicate datasets (across 12 experimental conditions) passed the Shapiro-Wilk and Kolmogorov-Smirnov normality tests. These results indicate we cannot reject the null hypothesis that the 76 datasets do not follow a normal or Gaussian distribution. It is also important to clarify that the 4-7 biological replicate trials conducted for each of the 12 experimental conditions (3 solute levels x 2 RH conditions x 2 phage concentrations) do not correspond to 4-7 individual droplet size measurements per size fraction within each condition. As shown in Table S37, the four trials conducted for 1.5% w/v solute content at 15% RH resulted in an average recovery of 102 - 585 viable pathogen-laden droplets. Therefore, given the high droplet counts measured per condition across the 4-7 independent trials, a parametric statistical analysis should provide a reliable statistical output.

**Table S37. Viable pathogen-laden droplet counts generated 1×10$^5$ PFU/mL equilibrated at 15± 5% RH**

| $d_{eq}$ size range (µm) | 1.5% w/v | | | 3% w/v | | | 6% w/v | | |
|---|---|---|---|---|---|---|---|---|---|
| | Mean | SD | N | Mean | SD | N | Mean | SD | N |
| ≥18.65 to ≥20.28 | 172 | 72 | 4 | 180 | 66 | 6 | 132 | 67 | 5 |
| 12.52 to 20.28 | 377 | 158 | 4 | 273 | 101 | 6 | 128 | 58 | 5 |
| 8.79 to 13.62 | 511 | 81 | 4 | 389 | 101 | 6 | 231 | 98 | 5 |
| 5.59 to 9.56 | 585 | 122 | 4 | 450 | 110 | 6 | 273 | 86 | 5 |
| 2.93 to 6.09 | 349 | 146 | 4 | 312 | 65 | 6 | 245 | 83 | 5 |
| 1.73 to 3.19 | 102 | 46 | 4 | 64 | 27 | 6 | 54 | 26 | 5 |

**Table S38. Viable pathogen-laden droplet counts generated 1×10$^5$ PFU/mL equilibrated at 5± 5% RH**

| $d_{eq}$ size range (µm) | 1.5% w/v | | | 3% w/v | | | 6% w/v | | |
|---|---|---|---|---|---|---|---|---|---|
| | Mean | SD | N | Mean | SD | N | Mean | SD | N |
| ≥18.65 to ≥20.28 | 103 | 33 | 7 | 139 | 51 | 7 | 132 | 52 | 7 |
| 12.52 to 20.28 | 223 | 75 | 7 | 212 | 67 | 7 | 114 | 50 | 7 |
| 8.79 to 13.62 | 349 | 141 | 7 | 311 | 107 | 7 | 171 | 72 | 7 |
| 5.59 to 9.56 | 356 | 140 | 7 | 325 | 86 | 7 | 205 | 57 | 7 |
| 2.93 to 6.09 | 226 | 79 | 7 | 238 | 88 | 7 | 177 | 58 | 7 |
| 1.73 to 3.19 | 32 | 17 | 7 | 40 | 18 | 7 | 43 | 18 | 7 |

**Table S39. Viable pathogen-laden droplet counts generated 5×10$^5$ PFU/mL equilibrated at 15± 5% RH**

| $d_{eq}$ size range (µm) | 1.5% w/v | | | 3% w/v | | | 6% w/v | | |
|---|---|---|---|---|---|---|---|---|---|
| | Mean | SD | N | Mean | SD | N | Mean | SD | N |
| ≥18.65 to ≥20.28 | 391 | 29 | 4 | 479 | 88 | 5 | 461 | 67 | 6 |
| 12.52 to 20.28 | 1023 | 250 | 4 | 917 | 375 | 5 | 505 | 144 | 6 |
| 8.79 to 13.62 | 1505 | 502 | 4 | 1585 | 506 | 5 | 1050 | 314 | 6 |
| 5.59 to 9.56 | 1531 | 630 | 4 | 2017 | 768 | 5 | 1544 | 371 | 6 |
| 2.93 to 6.09 | 918 | 662 | 4 | 1284 | 623 | 5 | 1291 | 485 | 6 |
| 1.73 to 3.19 | 197 | 102 | 4 | 250 | 143 | 5 | 460 | 431 | 6 |

**Table S40. Viable pathogen-laden droplet counts generated 5×10$^5$ PFU/mL equilibrated at 50± 5% RH**

| $d_{eq}$ size range (µm) | 1.5% w/v | | | 3% w/v | | | 6% w/v | | |
|---|---|---|---|---|---|---|---|---|---|
| | Mean | SD | N | Mean | SD | N | Mean | SD | N |
| ≥18.65 to ≥20.28 | 375 | 123 | 5 | 517 | 40 | 5 | 654 | 136 | 5 |
| 12.52 to 20.28 | 1122 | 648 | 5 | 794 | 178 | 5 | 496 | 114 | 5 |
| 8.79 to 13.62 | 1685 | 683 | 5 | 1314 | 464 | 5 | 782 | 202 | 5 |
| 5.59 to 9.56 | 1653 | 618 | 5 | 1599 | 421 | 5 | 1059 | 183 | 5 |
| 2.93 to 6.09 | 1323 | 457 | 5 | 1274 | 126 | 5 | 1135 | 451 | 5 |
| 1.73 to 3.19 | 219 | 72 | 5 | 228 | 74 | 5 | 231 | 76 | 5 |

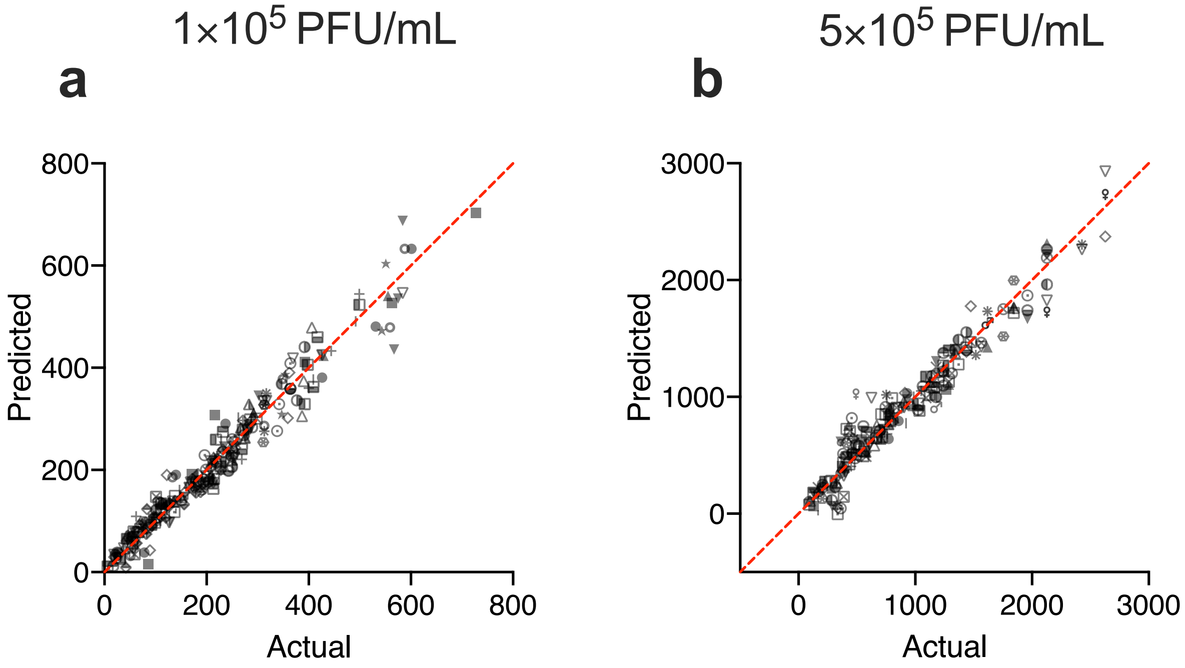


**Figure S4.** Normal QQ plots for Shapiro-Wilk and Kolmogorov-Smirnov normality tests.

### SECTION S10. TWO-WAY ANOVA FOLLOWED BY TUKEY'S MULTIPLE COMPARISON TEST RESULTS FOR RECOVERED COUNTS OF VIABLE PATHOGEN-LADEN DROPLETS COLLECTED BELOW EACH IMPACTOR STAGE

**Table S41. $1\times10^5$ PFU/mL - Viable pathogen-laden droplet counts collected below the first impactor stage**

| Tukey's multiple comparisons test | Summary | Adjusted P Value |
|---|---|---|
| [15]:1.5 vs. [15]:3 | ns | >0.9999 |
| [15]:1.5 vs. [15]:6 | ns | 0.8876 |
| [15]:1.5 vs. [50]:1.5 | ns | 0.3802 |
| [15]:1.5 vs. [50]:3 | ns | 0.9316 |
| [15]:1.5 vs. [50]:6 | ns | 0.8534 |
| [15]:3 vs. [15]:6 | ns | 0.7166 |
| [15]:3 vs. [50]:1.5 | ns | 0.1652 |
| [15]:3 vs. [50]:3 | ns | 0.7770 |
| [15]:3 vs. [50]:6 | ns | 0.6361 |
| [15]:6 vs. [50]:1.5 | ns | 0.9480 |
| [15]:6 vs. [50]:3 | ns | >0.9999 |
| [15]:6 vs. [50]:6 | ns | >0.9999 |
| [50]:1.5 vs. [50]:3 | ns | 0.8298 |
| [50]:1.5 vs. [50]:6 | ns | 0.9274 |
| [50]:3 vs. [50]:6 | ns | 0.9999 |

**Table S42. 1×10$^5$ PFU/mL - Viable pathogen-laden droplet counts collected below the second impactor stage**

| Tukey's multiple comparisons test | Summary | Adjusted P Value |
|---|---|---|
| [15]:1.5 vs. [15]:3 | ns | 0.4199 |
| [15]:1.5 vs. [15]:6 | ** | 0.0016 |
| [15]:1.5 vs. [50]:1.5 | ns | 0.0676 |
| [15]:1.5 vs. [50]:3 | * | 0.0413 |
| [15]:1.5 vs. [50]:6 | *** | 0.0003 |
| [15]:3 vs. [15]:6 | ns | 0.0779 |
| [15]:3 vs. [50]:1.5 | ns | 0.8907 |
| [15]:3 vs. [50]:3 | ns | 0.7760 |
| [15]:3 vs. [50]:6 | * | 0.0218 |
| [15]:6 vs. [50]:1.5 | ns | 0.4074 |
| [15]:6 vs. [50]:3 | ns | 0.5478 |
| [15]:6 vs. [50]:6 | ns | 0.9997 |
| [50]:1.5 vs. [50]:3 | ns | 0.9998 |
| [50]:1.5 vs. [50]:6 | ns | 0.1805 |
| [50]:3 vs. [50]:6 | ns | 0.2822 |

**Table S43. 1×10$^5$ PFU/mL - Viable pathogen-laden droplet counts collected below the third impactor stage**

| Tukey's multiple comparisons test | Summary | Adjusted P Value |
|---|---|---|
| [15]:1.5 vs. [15]:3 | ns | 0.4710 |
| [15]:1.5 vs. [15]:6 | ** | 0.0048 |
| [15]:1.5 vs. [50]:1.5 | ns | 0.1639 |
| [15]:1.5 vs. [50]:3 | * | 0.0484 |
| [15]:1.5 vs. [50]:6 | *** | 0.0002 |
| [15]:3 vs. [15]:6 | ns | 0.1600 |
| [15]:3 vs. [50]:1.5 | ns | 0.9828 |
| [15]:3 vs. [50]:3 | ns | 0.7634 |
| [15]:3 vs. [50]:6 | ** | 0.0092 |
| [15]:6 vs. [50]:1.5 | ns | 0.4070 |
| [15]:6 vs. [50]:3 | ns | 0.7808 |
| [15]:6 vs. [50]:6 | ns | 0.9200 |
| [50]:1.5 vs. [50]:3 | ns | 0.9827 |
| [50]:1.5 vs. [50]:6 | * | 0.0359 |
| [50]:3 vs. [50]:6 | ns | 0.1549 |

**Table S44. 1×10$^5$ PFU/mL - Viable pathogen-laden droplet counts collected below the fourth impactor stage**

| Tukey's multiple comparisons test | Summary | Adjusted P Value |
|---|---|---|
| [15]:1.5 vs. [15]:3 | ns | 0.3443 |
| [15]:1.5 vs. [15]:6 | ** | 0.0011 |
| [15]:1.5 vs. [50]:1.5 | * | 0.0143 |
| [15]:1.5 vs. [50]:3 | ** | 0.0042 |
| [15]:1.5 vs. [50]:6 | **** | <0.0001 |
| [15]:3 vs. [15]:6 | ns | 0.0758 |
| [15]:3 vs. [50]:1.5 | ns | 0.5747 |
| [15]:3 vs. [50]:3 | ns | 0.2726 |
| [15]:3 vs. [50]:6 | ** | 0.0022 |
| [15]:6 vs. [50]:1.5 | ns | 0.7354 |
| [15]:6 vs. [50]:3 | ns | 0.9504 |
| [15]:6 vs. [50]:6 | ns | 0.8703 |
| [50]:1.5 vs. [50]:3 | ns | 0.9927 |
| [50]:1.5 vs. [50]:6 | ns | 0.0963 |
| [50]:3 vs. [50]:6 | ns | 0.2769 |

**Table S45. 1×10$^5$ PFU/mL - Viable pathogen-laden droplet counts collected below the fifth impactor stage**

| Tukey's multiple comparisons test | Summary | Adjusted P Value |
|---|---|---|
| [15]:1.5 vs. [15]:3 | ns | 0.9850 |
| [15]:1.5 vs. [15]:6 | ns | 0.4743 |
| [15]:1.5 vs. [50]:1.5 | ns | 0.2212 |
| [15]:1.5 vs. [50]:3 | ns | 0.3230 |
| [15]:1.5 vs. [50]:6 | * | 0.0326 |
| [15]:3 vs. [15]:6 | ns | 0.7814 |
| [15]:3 vs. [50]:1.5 | ns | 0.4584 |
| [15]:3 vs. [50]:3 | ns | 0.6172 |
| [15]:3 vs. [50]:6 | ns | 0.0733 |
| [15]:6 vs. [50]:1.5 | ns | 0.9986 |
| [15]:6 vs. [50]:3 | ns | >0.9999 |
| [15]:6 vs. [50]:6 | ns | 0.7387 |
| [50]:1.5 vs. [50]:3 | ns | 0.9998 |
| [50]:1.5 vs. [50]:6 | ns | 0.8879 |
| [50]:3 vs. [50]:6 | ns | 0.7601 |

**Table S46. 1×10$^5$ PFU/mL - Viable pathogen-laden droplet counts collected below the sixth impactor stage**

| Tukey's multiple comparisons test | Summary | Adjusted P Value |
|---|---|---|
| [15]:1.5 vs. [15]:3 | ns | 0.2007 |
| [15]:1.5 vs. [15]:6 | ns | 0.0753 |
| [15]:1.5 vs. [50]:1.5 | ** | 0.0013 |
| [15]:1.5 vs. [50]:3 | ** | 0.0057 |
| [15]:1.5 vs. [50]:6 | ** | 0.0092 |
| [15]:3 vs. [15]:6 | ns | 0.9866 |
| [15]:3 vs. [50]:1.5 | ns | 0.2232 |
| [15]:3 vs. [50]:3 | ns | 0.5570 |
| [15]:3 vs. [50]:6 | ns | 0.6851 |
| [15]:6 vs. [50]:1.5 | ns | 0.6514 |
| [15]:6 vs. [50]:3 | ns | 0.9363 |
| [15]:6 vs. [50]:6 | ns | 0.9764 |
| [50]:1.5 vs. [50]:3 | ns | 0.9855 |
| [50]:1.5 vs. [50]:6 | ns | 0.9508 |
| [50]:3 vs. [50]:6 | ns | >0.9999 |

**Table S47. 5×10$^5$ PFU/mL - Viable pathogen-laden droplet counts collected below the first impactor stage**

| Tukey's multiple comparisons test | Summary | Adjusted P Value |
|---|---|---|
| [15]:1.5 vs. [15]:3 | ns | 0.7059 |
| [15]:1.5 vs. [15]:6 | ns | 0.8339 |
| [15]:1.5 vs. [50]:1.5 | ns | 0.9998 |
| [15]:1.5 vs. [50]:3 | ns | 0.3385 |
| [15]:1.5 vs. [50]:6 | ** | 0.0029 |
| [15]:3 vs. [15]:6 | ns | 0.9995 |
| [15]:3 vs. [50]:1.5 | ns | 0.4786 |
| [15]:3 vs. [50]:3 | ns | 0.9841 |
| [15]:3 vs. [50]:6 | ns | 0.0546 |
| [15]:6 vs. [50]:1.5 | ns | 0.6214 |
| [15]:6 vs. [50]:3 | ns | 0.9101 |
| [15]:6 vs. [50]:6 | * | 0.0201 |
| [50]:1.5 vs. [50]:3 | ns | 0.1712 |
| [50]:1.5 vs. [50]:6 | *** | 0.0008 |
| [50]:3 vs. [50]:6 | ns | 0.2011 |

**Table S48. 5×10$^5$ PFU/mL - Viable pathogen-laden droplet counts collected below the second impactor stage**

| Tukey's multiple comparisons test | Summary | Adjusted P Value |
|---|---|---|
| [15]:1.5 vs. [15]:3 | ns | 0.9969 |
| [15]:1.5 vs. [15]:6 | ns | 0.2003 |
| [15]:1.5 vs. [50]:1.5 | ns | 0.9976 |
| [15]:1.5 vs. [50]:3 | ns | 0.9088 |
| [15]:1.5 vs. [50]:6 | ns | 0.2194 |
| [15]:3 vs. [15]:6 | ns | 0.3570 |
| [15]:3 vs. [50]:1.5 | ns | 0.9250 |
| [15]:3 vs. [50]:3 | ns | 0.9915 |
| [15]:3 vs. [50]:6 | ns | 0.3815 |
| [15]:6 vs. [50]:1.5 | ns | 0.0566 |
| [15]:6 vs. [50]:3 | ns | 0.7140 |
| [15]:6 vs. [50]:6 | ns | >0.9999 |
| [50]:1.5 vs. [50]:3 | ns | 0.6412 |
| [50]:1.5 vs. [50]:6 | ns | 0.0684 |
| [50]:3 vs. [50]:6 | ns | 0.7260 |

**Table S49. 5×10$^5$ PFU/mL - Viable pathogen-laden droplet counts collected below the third impactor stage**

| Tukey's multiple comparisons test | Summary | Adjusted P Value |
|---|---|---|
| [15]:1.5 vs. [15]:3 | ns | 0.9998 |
| [15]:1.5 vs. [15]:6 | ns | 0.6555 |
| [15]:1.5 vs. [50]:1.5 | ns | 0.9915 |
| [15]:1.5 vs. [50]:3 | ns | 0.9890 |
| [15]:1.5 vs. [50]:6 | ns | 0.2241 |
| [15]:3 vs. [15]:6 | ns | 0.4232 |
| [15]:3 vs. [50]:1.5 | ns | 0.9993 |
| [15]:3 vs. [50]:3 | ns | 0.9366 |
| [15]:3 vs. [50]:6 | ns | 0.1044 |
| [15]:6 vs. [50]:1.5 | ns | 0.2479 |
| [15]:6 vs. [50]:3 | ns | 0.9317 |
| [15]:6 vs. [50]:6 | ns | 0.9283 |
| [50]:1.5 vs. [50]:3 | ns | 0.8004 |
| [50]:1.5 vs. [50]:6 | ns | 0.0516 |
| [50]:3 vs. [50]:6 | ns | 0.4769 |

**Table S50. 5×10$^5$ PFU/mL - Viable pathogen-laden droplet counts collected below the fourth impactor stage**

| Tukey's multiple comparisons test | Summary | Adjusted P Value |
|---|---|---|
| [15]:1.5 vs. [15]:3 | ns | 0.7380 |
| [15]:1.5 vs. [15]:6 | ns | >0.9999 |
| [15]:1.5 vs. [50]:1.5 | ns | 0.9993 |
| [15]:1.5 vs. [50]:3 | ns | >0.9999 |
| [15]:1.5 vs. [50]:6 | ns | 0.7597 |
| [15]:3 vs. [15]:6 | ns | 0.6747 |
| [15]:3 vs. [50]:1.5 | ns | 0.8780 |
| [15]:3 vs. [50]:3 | ns | 0.8032 |
| [15]:3 vs. [50]:6 | ns | 0.0770 |
| [15]:6 vs. [50]:1.5 | ns | 0.9993 |
| [15]:6 vs. [50]:3 | ns | >0.9999 |
| [15]:6 vs. [50]:6 | ns | 0.6511 |
| [50]:1.5 vs. [50]:3 | ns | >0.9999 |
| [50]:1.5 vs. [50]:6 | ns | 0.4901 |
| [50]:3 vs. [50]:6 | ns | 0.5897 |

**Table S51. 5×10^5 PFU/mL - Viable pathogen-laden droplet counts collected below the fifth impactor stage**

| Tukey's multiple comparisons test | Summary | Adjusted P Value |
|---|---|---|
| [15]:1.5 vs. [15]:3 | ns | 0.8716 |
| [15]:1.5 vs. [15]:6 | ns | 0.8425 |
| [15]:1.5 vs. [50]:1.5 | ns | 0.8167 |
| [15]:1.5 vs. [50]:3 | ns | 0.8831 |
| [15]:1.5 vs. [50]:6 | ns | 0.9846 |
| [15]:3 vs. [15]:6 | ns | >0.9999 |
| [15]:3 vs. [50]:1.5 | ns | >0.9999 |
| [15]:3 vs. [50]:3 | ns | >0.9999 |
| [15]:3 vs. [50]:6 | ns | 0.9965 |
| [15]:6 vs. [50]:1.5 | ns | >0.9999 |
| [15]:6 vs. [50]:3 | ns | >0.9999 |
| [15]:6 vs. [50]:6 | ns | 0.9946 |
| [50]:1.5 vs. [50]:3 | ns | >0.9999 |
| [50]:1.5 vs. [50]:6 | ns | 0.9895 |
| [50]:3 vs. [50]:6 | ns | 0.9974 |

**Table S52. 5×10[5] PFU/mL - Viable pathogen-laden droplet counts collected below the sixth impactor stage**

| Tukey's multiple comparisons test | Summary | Adjusted P Value |
|---|---|---|
| [15]:1.5 vs. [15]:3 | ns | 0.9990 |
| [15]:1.5 vs. [15]:6 | ns | 0.4267 |
| [15]:1.5 vs. [50]:1.5 | ns | >0.9999 |
| [15]:1.5 vs. [50]:3 | ns | >0.9999 |
| [15]:1.5 vs. [50]:6 | ns | 0.9999 |
| [15]:3 vs. [15]:6 | ns | 0.5966 |
| [15]:3 vs. [50]:1.5 | ns | >0.9999 |
| [15]:3 vs. [50]:3 | ns | >0.9999 |
| [15]:3 vs. [50]:6 | ns | >0.9999 |
| [15]:6 vs. [50]:1.5 | ns | 0.4523 |
| [15]:6 vs. [50]:3 | ns | 0.4941 |
| [15]:6 vs. [50]:6 | ns | 0.5089 |
| [50]:1.5 vs. [50]:3 | ns | >0.9999 |
| [50]:1.5 vs. [50]:6 | ns | >0.9999 |
| [50]:3 vs. [50]:6 | ns | >0.9999 |

### SECTION S11. TWO-WAY ANOVA FOLLOWED BY TUKEY'S MULTIPLE COMPARISON TEST RESULTS FOR RECOVERED COUNTS OF VIABLE PATHOGEN-LADEN DROPLETS COLLECTED BELOW ALL IMPACTOR STAGES

**Table S53. 1×$10^5$ PFU/mL - Viable pathogen-laden droplet counts collected below all impactor stages**

| Tukey's multiple comparisons test | Summary | Adjusted P Value |
|---|---|---|
| 15±5:1.5 vs. 15±5:3 | ns | 0.4423 |
| 15±5:1.5 vs. 15±5:6 | ** | 0.0019 |
| 15±5:1.5 vs. 50±5:1.5 | * | 0.0125 |
| 15±5:1.5 vs. 50±5:3 | ** | 0.0095 |
| 15±5:1.5 vs. 50±5:6 | **** | <0.0001 |
| 15±5:3 vs. 15±5:6 | ns | 0.0837 |
| 15±5:3 vs. 50±5:1.5 | ns | 0.4132 |
| 15±5:3 vs. 50±5:3 | ns | 0.3472 |
| 15±5:3 vs. 50±5:6 | ** | 0.0030 |
| 15±5:6 vs. 50±5:1.5 | ns | 0.8839 |
| 15±5:6 vs. 50±5:3 | ns | 0.9240 |
| 15±5:6 vs. 50±5:6 | ns | 0.8929 |
| 50±5:1.5 vs. 50±5:3 | ns | >0.9999 |
| 50±5:1.5 vs. 50±5:6 | ns | 0.2066 |
| 50±5:3 vs. 50±5:6 | ns | 0.2569 |

**Table S54. $5×10^5$ PFU/mL - Viable pathogen-laden droplet counts collected below all impactor stages**

| Tukey's multiple comparisons test | Summary | Adjusted P Value |
|---|---|---|
| 15±5:1.5 vs. 15±5:3 | ns | 0.9659 |
| 15±5:1.5 vs. 15±5:6 | ns | >0.9999 |
| 15±5:1.5 vs. 50±5:1.5 | ns | 0.9840 |
| 15±5:1.5 vs. 50±5:3 | ns | >0.9999 |
| 15±5:1.5 vs. 50±5:6 | ns | 0.9164 |
| 15±5:3 vs. 15±5:6 | ns | 0.8725 |
| 15±5:3 vs. 50±5:1.5 | ns | >0.9999 |
| 15±5:3 vs. 50±5:3 | ns | 0.9800 |
| 15±5:3 vs. 50±5:6 | ns | 0.4303 |
| 15±5:6 vs. 50±5:1.5 | ns | 0.9231 |
| 15±5:6 vs. 50±5:3 | ns | 0.9989 |
| 15±5:6 vs. 50±5:6 | ns | 0.9508 |
| 50±5:1.5 vs. 50±5:3 | ns | 0.9923 |
| 50±5:1.5 vs. 50±5:6 | ns | 0.5093 |
| 50±5:3 vs. 50±5:6 | ns | 0.8365 |

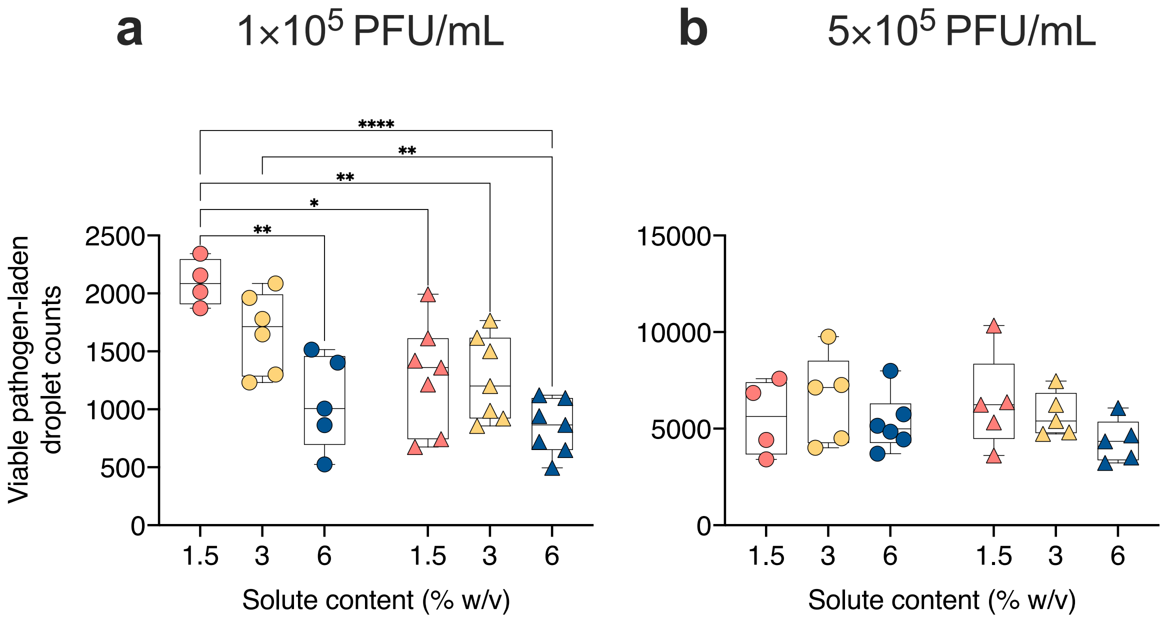


**Figure S5. Effect of solute content, relative humidity, and phage concentration on recovered counts of viable pathogen-laden droplets collected below all impactor stages.** Phi6 bacteriophage suspensions with concentrations of (**a**) $1\times10^5$ PFU/mL and (**b**) $5\times10^5$ PFU/mL were prepared in 1.5% (**coral**), 3% (**yellow**), and 6% (**blue**) w/v peptone water solutions and aerosolized into 15±5% (**circles**) and 50±5% (**triangles**) relative humidity conditions at 24±1.6°C. Graphs present coincidence-corrected viable droplet counts collected below all impactor stages with an initial size range of 1.73 to ≥20.28 μm. Significance determined using two-way ANOVA followed by Tukey's multiple comparisons tests defined as * - $p<0.05$, ** - $p<0.01$, **** - $p<0.0001$.

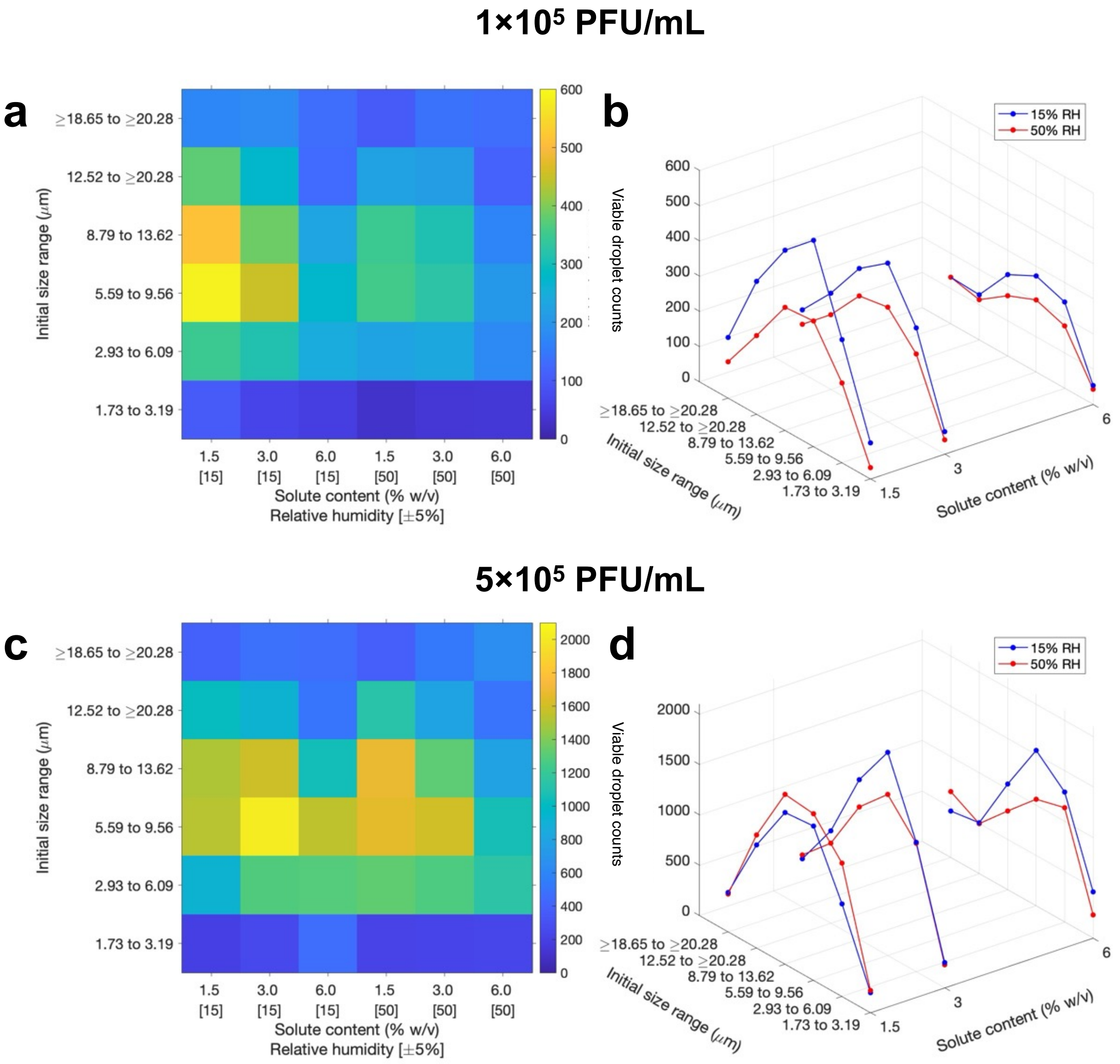


**Figure S6. Initial size range influences solute-, relative humidity-, and phage concentration-dependent changes in viable pathogen-laden droplet counts.** (**a,c**) Intensity maps and (**b,d**) 3D plots present viable pathogen-laden droplet counts with initial size range of 1.73 to ≥20.28 µm generated from Phi6 suspensions with concentrations of (**a,b**) $1\times10^5$ PFU/mL and (**c,d**) $5\times10^5$ PFU/mL prepared in 1.5%, 3%, and 6% w/v peptone water solutions and aerosolized into 15±5% and 50±5% relative humidity conditions at 24±1.6°C.
